# Placental LBX1 dysregulation drives ribosomal dysfunction and unfolded protein response in spontaneous preterm birth

**DOI:** 10.64898/2026.09.11.26362823

**Authors:** Ankit Biswas, Naman Kharbanda, Krishna Singh Bisht, Pragya Tailor, Sandhini Saha, Arundhati Tiwari, Mimansa Sharma, Ramchandran Thiruvengadam, Nitya Wadhwa, Dinakar M Salunke, Shinjini Bhatnagar, GARBH-Ini study team, Pallavi Kshetrapal, Tushar Kanti Maiti

## Abstract

Spontaneous preterm birth (sPTB) affects ∼9.9% of pregnancies worldwide and is driven in part by placental dysfunction, yet the signalling events linking placental dysregulation to spontaneous prematurity remain incompletely defined. Using a cross-sectional design, we profiled 70 placentae from spontaneous term and preterm birth delivery to construct protein co-expression networks and identified 13 hub proteins associated with sPTB. Quantification of placental circulatory proteins in an independent nested case-control set of 53 maternal plasma samples highlighted elevated levels of the transcription factor ladybird homeobox protein 1 (LBX1) in maternal circulation from sPTB cases. Elevated LBX1 in trophoblast cells undergo KPNB1 mediated nuclear import, where it deregulates ribosomal biogenesis and perturbs subsequent ribosomal protein homeostasis, contributing to the accumulation of newly synthesised proteins. Accumulation of these nascent proteins triggers unfolded protein response which may lead to adverse pregnancy outcome. These findings implicate LBX1-mediated ribosomal disruption and activation of unfolded protein response in trophoblasts as a placental mechanism contributing to sPTB.

**Graphical Abstract:** 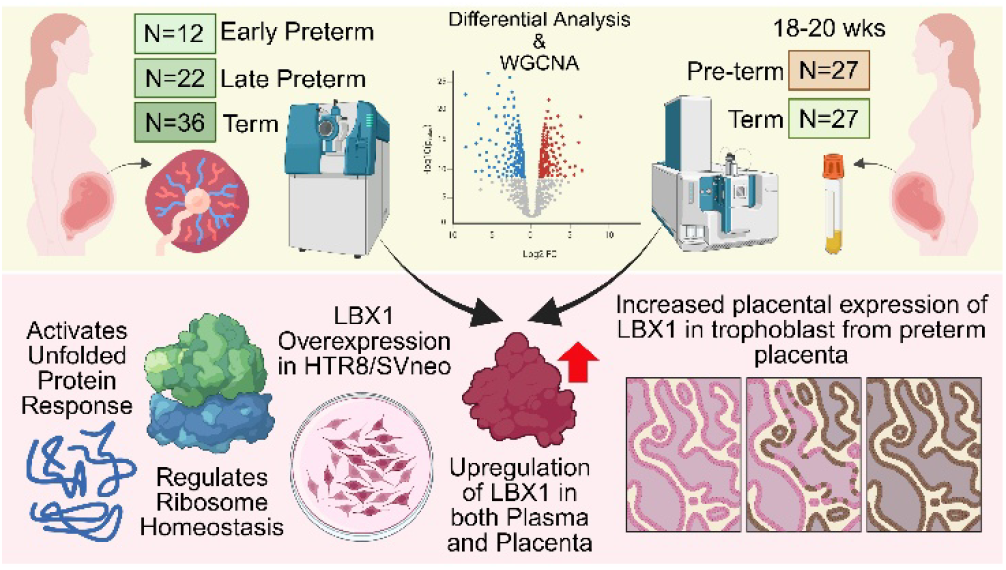

## 1. Introduction

Preterm birth (PTB) refers to the delivery of neonates before the completion of 37 weeks of gestation. With 13.4 million babies born premature in 2020 alone, PTB constitutes a considerable share of pregnancy-related complications annually worldwide^1,2^. Based on the period of gestation (POG), early preterm (less than 34 weeks) and late preterm (34-37 weeks) babies are at higher risk of adverse childhood health outcomes as compared to the term babies (more than 37 weeks). Thus, PTB has far-reaching social consequences with increased predisposition to lifelong morbidities such as developmental challenges, health complications ranging from pulmonary, cardiac to neurological and mortality figures standing at 1 million children every year^2,3^. While pregnancy-related complications and existing medical conditions warrant expedited iatrogenic delivery, 45-50% of premature births are of an idiopathic nature or spontaneous (sPTB)^4^. Pathologies such as premature rupture of membrane (PROM) and spontaneous preterm labour also result in sPTB. The partially understood factors contributing to sPTB include, but are not limited to, maternal age, pre-existing medical conditions and lifestyle factors like smoking, alcohol consumption and environmental factors^5^. The complex and elusive etiology, coupled with associated mortality, makes the prevention and management of the condition challenging. Thus, preterm birth continues to remain a major global health problem and a subject of continued research to refine the prevention, prediction and management of this constellation of complications.

Placenta is a transient organ that acts as a conduit at the junction of the feto-maternal system to facilitate the exchange of gasses, nutrients and waste. Beyond its role of providing nutrients, the placenta also mediates endocrine and immune functions for proper growth and development of the fetus^6^. Once the fetus matures, the organ promotes parturition by initiating labour. Studies report correlation between sPTB and placental insufficiency^7^. Thus, identification of differential molecular signature of the sPTB placenta may provide valuable information related to the pathology. Previous proteomic studies carried out with similar objectives were conducted on relatively small sample sizes using maternal sera, saliva or high vaginal fluid from age-matched term and pre-term delivered mothers. However, mechanistic studies conducted on bodily fluids provide only limited information about the biomolecules leaching from organs and thus yield a skewed picture of cellular signalling events. Moreover, multiple omics studies undertaken on the placental tissue samples have not considered the placental age and associated signalling dynamics, as a false positive detection for altered signalling in sPTB. Therefore, we employed a multi-stage cross sectional study design to select independent participants at multiple gestational ages across the POG. Placental samples obtained at these different gestational time points were used to characterize placental molecular signatures as a continuous function of gestational age, thereby defining the trajectory of normal placental aging. These age-related trajectories were subsequently compared with placental signatures associated with sPTB to distinguish physiological placental aging from sPTB-associated molecular alterations. Thus, the identified alterations identified were further monitored in maternal plasma samples collected from participants at 18-20 weeks of gestation and selected using a nested case control study design.

In this study, we employed data-independent acquisition-mass spectrometry (DIA-MS) based quantitation of 70 clinical placental samples for identification and quantitation of differential protein expression in sPTB. Since the lifelong morbidity and mortality of the neonates is greatly influenced by the gestational age at the time of delivery, the study was trichotomized into EPTB (<34 weeks), LPTB (34-37 weeks), and TB (>37 weeks) groups. SWATH-acquisition revealed 73 and 31 differentially expressed proteins (DEPs) in EPTB vs TB and LPTB vs TB comparison groups, respectively. Protein co-expression network analysis identified five co-expressing modules; two of which were significantly correlated with the comparison groups. The overlap with DEPs yielded thirteen co-expressing proteins that were further validated in clinical plasma samples to highlight five upregulated proteins that are thrombospondin 1 (THBS1), calponin 1 (CNN1), ladybird homeobox protein 1 (LBX1), TGF-beta induced protein (TGFB1), and creatine kinase B (CKB). Interestingly, the LBX1 protein showed elevated expression in placenta and the maternal plasma. The presence of LBX1was validated in placental trophoblast cells. Further exploration utilizing differential proteomics and interactomics revealed a non-canonical role of LBX1 in ribosomal homeostasis. Our findings report that LBX1 regulates ribosomal functions to elevate the nascent protein synthesis. These accumulated nascent proteins in trophoblast cells trigger the unfolded protein response (UPR). Thus, LBX1 over-expression in extravillous trophoblast cells can lead to ribosomal dysfunction, facilitating the UPR mediated placental damage during spontaneous preterm birth.

## Methods

### 2.1 Study population and clinical definitions

The interdisciplinary Group for Advanced Research on Birth outcomes-DBT India Initiative (GARBH-Ini) is an ongoing prospective observational hospital-based pregnancy cohort in the North of India with the primary aim to generate a risk-prediction algorithm for PTB based on multidimensional risk factors assessed during pregnancy. Detailed objectives and cohort information can be assessed from our previous study.^8^ The enrolled participants represent a mix of semi-urban and rural populations of Haryana. Women before 20 weeks of pregnancy are enrolled and followed up to five times until delivery and once within 6 months postpartum. A diverse range of biological specimens are collected, with ultrasound scans conducted at regular intervals according to the study protocol. Ethical approval for this study was obtained from the Institutional Ethics Committees of all the collaborating institutions. For the discovery study using placental samples, we employed a cross-sectional study design to capture molecular signals from term and preterm delivered mothers, treating POG as a continuous variable. Samples were selected across a broad range of POG, recognizing the fact that the placenta is accessible only after delivery. The defined universe yielded 1502 eligible participants with placental samples available in the cohort. The participants with singleton, normal vaginal delivery are included, whereas mothers with pregnancy-induced hypertension, preeclampsia, eclampsia, gestational diabetes mellitus, fetal anomaly, malaria, no information for neonatal gender, previous history of preterm (PT), smoking, alcohol and steroid consumption were excluded. The remaining 762 participants are screened for early-preterm (less than 34 weeks), late-preterm (35-37 weeks) and term (37-39 weeks) subgroups according to recent ACOG guidelines^9^ to randomly select 12 early preterm, 22 late-preterm and 36 term participants from the cohort. The early and late preterm are considered cases, while the term delivered mothers are used as controls (**Supplementary Figure 1A**).

In the next phase the circulatory placental signatures were tracked in second trimester maternal plasma employing a nested case-control study design. The defined universe with inclusion criteria of live, singleton, spontaneous birth with availability of plasma samples at 18-20 and 26-28 weeks of gestation selected 887 participants. The discovery phase matching exclusion criteria are utilised along with matching criteria for delivery date (within the same month), parity, and sex of the newborn to randomly select 26 cases and 27 control participants from the cohort. Mothers who delivered preterm (less than 35) were considered as case and mothers delivering term (more than 39) are denoted as controls (**Supplementary Figure 1B**).

### 2.2 Placental collection

Placental biopsies were collected within thirty minutes of delivery. The tissue punches (10 mm) from four designated placental quadrants were taken and then rinsed in ice cold PBS twice. The wet tissue was dabbed to remove extra liquid and immediately flash freezed. These placental biopsies were then archived at −80 °C at the institutional biorepository until further processing. For immuno-histochemical analysis formalin fixed paraffin embedded blocks were prepared using fresh placental tissues. Briefly, a 1-inch X 1-inch region of entire placental thickness were incised using scalpel and scissors, followed by sterile ice-cold PBS rinse. The sampled tissues were transferred in a 50 mL falcon containing 10% neutral buffered formalin (HT401128, Sigma) for fixation. The formalin fixed samples were stored at room temperature, with media replacement in every 6 months, until it is ready for block embedding. The placental tissue blocks were embedded in paraffin and then stored at RT until immuno-histochemical slide preparation.

### 2.3 Placental protein extraction and tryptic digestion

The stored placental punches from four defined quadrants were washed in ice-cold PBS and 10 mg tissue from each quadrant were weighed and pooled together for protein extraction. The placental tissues were cryo-homogenized in liquid nitrogen and collected in RIPA (R0278, Sigma) lysis buffer added with 1X halt-protease and phosphatase inhibitor cocktail (78441, Thermo Scientific). Next, the lysates were sonicated and then centrifuged at 14,000 rpm for 45 min at 4 °C to collect the placental protein extract. These extracts were acetone (01408681, Wako) precipitated and again dissolved in 8M urea (219-00175, Wako) followed by protein quantification using BCA protein assay kit (23225, Pierce, Thermo Scientific). Next, 100 µg protein was aliquoted from each sample and reduced with 10 mM DL-Dithiothreitol (DTT) (D5545, Sigma) for 1 h at 56 °C followed by alkylation with 20 mM Iodoacetamide (IAA) (I1149, Sigma) in dark at room temperature for 1 hour. These reduced and alkylated proteins were then digested with sequencing grade trypsin (1:20 w/w in 50 mM ABC) (90057, Pierce, Thermo Scientific) at 37 °C for overnight. The reaction was stopped by drying the tryptic digest in a vacuum centrifuge, followed by desalting using Pierce C18 pipette tips (87784, Thermo scientific) and stored at −80 °C until ready for MS acquisition.

### 2.4 Data independent acquisition and processing

The digested and desalted peptides from each placenta sample (100 µg) were dissolved in solvent A (98% H2O and 2% acetonitrile with 0.1% formic acid) and spiked with iRT reagent (1816351, Biognosys) as per the manufacturer’s instructions. The tryptic digests were analyzed on a Triple TOF 5600+ (Sciex, Concord Canada) mass spectrometer coupled with ekspert nanoLC 425 (Eksigent, Dublin, CA, USA) system. 8 µl digests from each sample (10 µg of sample + 1 µl of iRT) were loaded onto ChromXP C18 CL (10 cm X 300 µm, 5 µm, 120 Å, Eksigent) trap column, in triplicate, with a flow rate of 10 µL/min for 10 minutes. The tryptic peptides were then eluted from ChromXP C18 (150 mm X 300 um, 3 µm, 100 Å, Eksigent) analytical column with a flow rate of 5 µL/min using a linear gradient from 2% solvent B (95% (v/v) acetonitrile with 0.1% (v/v) formic acid) to 90% solvent B in 78 minutes for a total run time of 87 minutes. A SWATH acquisition scheme for 60 overlapping variable size windows, covering 400-1250 Da mass range, were utilized with accumulation time of 11 msec and total cycle time of 1.4 sec.

The acquired SWATH-MS data was analyzed on Spectronaut Pulsar 14.9 (Biognosys). Briefly, a de novo spectral library was first prepared by searching the acquired .wiff files against the UniProtKB human protein database (20,214 entries, August 2017) with default search parameters. The library compiled 8,836 precursors and 7026 peptides from the entire run. The data were then searched against the generated library using default settings: RT prediction type was set to dynamic iRT, interference correction at MS2 level was enabled, and global TIC (total intensity count) was used for normalization of data, trypsin was selected as specific enzyme with maximum 2 missed cleavages. The Carbamidomethylation of cysteine (+57.021464 Da) were used for fixed modification whereas, methionine oxidation (+15.994915 Da), and N-terminal acetylation (+42.010565 Da) was selected for variable modifications. FDR was set to 1% at both peptide and precursor levels. The extracted-ion peak intensities of peptides were used for protein abundance calculation. These abundance values along with fold change and significance were extracted as report file, which was further used for quantitative placental proteome analysis. The SWATH data are available via ProteomeXchange with identifier PXD065403.

### 2.5 Protein co-expression network analysis

Protein co-expression network analysis was performed on R package WGCNA (v1.73) to identify the modules clustering as groups of highly correlated proteins in terms of their expression values. To build a scale-free network adjacency correlation between two genes p and q, it was calculated as, pq = power(S_pq_, β) = S_pq_(power)β, where S_pq_ = |cor(p, q)| is the gene co-expression similarity between two genes. For β calculation, independence and mean connectivity were tested using a gradient method (the power value ranging from 1 to 20). When the degree of independence was above 0.85, an appropriate power value was screened out.

Thus, normalized protein abundance data of all the 1550 protein groups present in 70 samples was exported in the form of a matrix from Spectronaut. Two samples with more than 30% missing values were excluded from subsequent analysis. The remaining missing values were then imputed using predictive mean matching (PMM) algorithm employing the MICE package (v3.17.0) in R, followed by log transformation. Next, an appropriate soft threshold power for scale-free topology was calculated and selected with the pickSoftThreshold function of the WGCNA package based on scale-free topology and mean connectivity parameters. A correlation matrix for all pairwise correlations of proteins across all samples was generated and then transformed into a weighted adjacency matrix with the selected soft threshold value of 7. Employing this adjacency matrix the Topological Overlap (TO) was measured. TO reflects a substantial pairwise measure of paired protein similarity based on their co-expression relation with the rest of the proteins in the network. Proteins were hierarchically clustered using topological overlap dissimilarity (1-TO) as the distance measure to generate a cluster dendrogram. Proteins with comparative co-expressions were grouped in modules using a dynamic tree-cutting algorithm based on the following major parameters: minimal module size = 50, deepSplit = 0, and merge cut height = 0.25. Next, the module eigengene (ME) was summarized by the first principal component of its representative matrix.

Module membership (kME) was determined by calculating Pearson correlation between individual protein and each module eigengene and their corresponding P Values. Subsequently, module–trait relationships were estimated using the correlation between MEs and clinical traits, which allowed efficient identification of the relevant modules. To evaluate the correlation strength, we calculated the module significance (MS), which is defined as the average absolute gene significance (GS) of all the genes involved in the module. The GS is measured as the log10 transformation of the P value (logP) in the linear regression between gene expression and clinical information.

### 2.6 Plasma collection

The venous blood samples were collected from mothers during the 18–20-week period of gestation in a sodium citrate-coated tube. The collected maternal blood was centrifuged at 2000 rpm for 20 min to separate the plasma. The isolated plasma was added with 1X halt-protease phosphatase inhibitor (78441, Thermo Scientific) and subsequently stored at −80 °C until sample retrieval was required.

### 2.7 Plasma protein digestion

The stored plasma samples were diluted 20-fold in 100 mM ammonium bi-carbonate (ABC) buffer and 20 µl were aliquoted from each sample. Proteins in the dilute plasma were reduced with 10 mM DTT at 56 °C for 30 mins and alkylated with 20 mM IAA, at RT for 1 h. The reduced and alkylated proteins were digested with MS-grade trypsin (90057, Pierce, Thermo Scientific), at 55 °C for 6 h. The digested protein samples were passed through Oasis HLB C18 cartridges (WAT094225, Waters) to remove residual salts and buffer ions. The peptides were then sequentially eluted in 80% and 100% acetonitrile, with 0.1% formic acid. Eluted samples were vacuum dried and stored at −80 °C until further use.

### 2.8 HR-MRM data acquisition and analysis

The targeted mass spectrometry was carried out on Sciex ZenoTOF 7600 instrument associated with a waters microscale LC system (ACQUITY UPLC M-Class System) in HR-MRM mode. Equivalent quantities of digested plasma peptides (1 µg) were placed into the Luna 5 µm C18 Micro trap column (20 mm X 0.3 mm, 100 Å, Phenomenex). The tryptic peptides were then resolved using a nanoEase M/Z HSS T3 analytical column (150 mm X 300 µm, 1.8 µm, 100 Å, Waters,) at a flow rate of 5 µL/min in a linear gradient of solvent B (100% (v/v) acetonitrile with 0.1% (v/v) formic acid) for 22 minutes, for a total run duration of 32 min. The most intense y and b ions of the targeted proteins were selected as precursors (transition) from the SRMatlas Database^10^ and SWATH data. The data was acquired in positive ion mode with ion source temperature of 200 °C and spray voltage of 5500 V. Scheduling of HR-MRM transitions was performed using accumulation time of 0.1 s and retention time tolerance of 20 s for each transition. Maximum scan time was set to 1.202 sec for 57 TOF-MS/MS scans in a single cycle, adding to 1597 cycles during total scan duration.

The raw HR-MRM data was further analyzed using MultiQuant integrated in Sciex OS (v 2.0.0.45330, Sciex). The MQ4 integration algorithm within MultiQuant was utilized for peak integration. All targeted spectral peaks were checked manually to ensure correct peak detection and accurate integration. The peak area of two transitions per precursor was traced for quantitation and proteins with at least ≥2 precursors were considered for comparison. The most intense precursors and its respective peptide were explored for differential regulation across all the 53 maternal plasma samples. These precursor intensity values of the respective proteins in all the samples were further used for downstream analysis. The HR-MRM data are available via ProteomeXchange with identifier PXD065014.

### 2.9 Cell culture, transfection and treatment

HTR8/SVneo, an immortalized first-trimester extravillous cells of trophoblastic lineage, were maintained in Roswell Park Memorial Institute (RPMI) 1640 medium (R6504, Sigma) which contains 2g/L sodium bicarbonate (40151UR-K05, SDFCL), 100 units/mL penicillin, 100 g/mL streptomycin (A001A, Himedia) and supplemented with 10% heat-inactivated fetal bovine serum (FBS) (A5256501, Gibco). The cells were kept in a CO_2_ incubator at 37 °C, with 5% CO_2_ and 85% relative humidity for optimal growth.

The cells were passaged 3 times post-revival before seeding in to a 6 well plate (3X10^5^ cells/well). The plates were incubated until the cells reaches suitable confluency (65%) for transfection with either plasmid or siRNA. For over-expression, mammalian expression vector (pcDNA3.1) containing flag-tag human LBX1 gene (OHu18208, GenScript), were transfected using lipofectamine 2000 (11668019, Thermo Scientific) following the manufacturer’s guidelines. Similarly, for transient knock down lipofectamine based transfection of SMARTpool ON-TARGETplus Human LBX1 siRNA (L-012289-00-0005, Horizon) was performed. Both the knock down and over-expression were confirmed using western blots. pcDNA3.1 empty vector and scrambled siRNA (SIC001, Sigma) were used in the respective control wells for over-expression and knock-down condition.

For, Importazole (IPZ) treatment the plasmid or siRNA transfected cells were kept till 24 hours, followed by replacing DMSO dissolved IPZ (10uM) (21491, Cayman chemicals) containing media in treated wells. Whereas the control wells were added with 0.1% DMS0 in complete RPMI media. The cells were further incubated for 24 hours and finally harvested in RIPA buffer for western blot analysis.

The nascent protein expression was visualized via puromycin incorporation assay, where 5 μg/ml puromycin (P8833, Sigma) were added to each well post 48 hours of transfection and incubated for 30 minutes. Then the cells were harvested for western blot analysis against anti-puromycin antibody (1:1000, A23031, Proteintech).

### 2.10 Proteomics sample preparation for transfected cells

The LBX1 over-expression and knock-down of HTR8/SVneo cells were confirmed after 48 hours of transfection followed by protein extraction in RIPA (R0278, Sigma) buffer containing 1X halt-protease and phosphatase inhibitor cocktail (78441, Thermo Scientific). Briefly, the cells lysates were sonicated and centrifuged to collect the supernatant containing total extracted protein. This protein extracts were acetone precipitated, dried and again redissolved in 8M urea, which was further diluted to 2M urea using 100 mM ammonium bicarbonate (ABC) (A6141, Sigma) buffer. The protein concentration was measured using BCA protein assay kit (23225, Pierce, Thermo Scientific) and 45 µg protein from each lysate were collected for subsequent reduction (10 mM DTT, 56 °C, 45 minutes) and alkylation (20 mM IAA, RT, 1 hour) steps. Next, the reduced and alkylated sample were trypsin digested using MS grade trypsin (90057, Pierce, Thermo Scientific) in 1:15 (enzyme: protein) ratio at 37 °C for overnight. After successful digestion the tryptic peptides were desalted using Oasis HLB C18 cartridges (WAT094225, Waters) and vacuum dried for storage at −80 °C until ready for MS data acquisition.

### 2.11 ZenoSWATH acquisition and data processing

The tryptic peptides were dissolved in solvent A (2% (vol/vol) acetonitrile, 0.1% (vol/vol) formic acid in water) and then profiled using Sciex ZenoTOF 7600 mass spectrometer coupled with waters microscale LC system (ACQUITY UPLC M-Class System). 500 ng digest from each samples were injected onto Luna 5µm C18 Micro trap column (20 mm X 0.3 mm, 100 Å, phenomenex) followed by peptide separation using a nanoEase M/Z HSS T3 analytical column (150 mm X 300 um, 1.8 µm, 100 Å, Waters,) for 22 minutes over a liner gradient of solvent B (100% (v/v) acetonitrile with 0.1% (v/v) formic acid). The flow rate was maintained at 5 µL/min for a total 32 minutes of runtime including washing and equilibration with solvent A at end. Triplicate data were acquired for each sample in a Zeno pulsing enabled SWATH mode with optimized settings for voltage, curtain gas, nebulizer gas, heater gas, and the source temperature. 65 overlapping variable windows were utilized to acquire the MS2 spectra (mass range) for 20 msec of accumulation period and 1.74 sec of cycle time.

A de-novo library was created with the acquired .wiff files in Spectronaut pulsar 19 (Biognosys) against the UniProtKB human protein database with isoforms (42,502 entries, December 2024). The library was then utilized to identify proteins and peptides from the acquired raw data with default settings enabled. Briefly, trypsin is selected as specific enzyme allowing only 2 missed cleavages with fixed modification of cysteine Carbamidomethylation (+57.021464 Da). While, variable modification of methionine oxidation (+15.994915 Da), and N-terminal acetylation (+42.010565 Da) was used for proteolytic peptide filtering. To ensure robust quantification dynamic iRT enabled RT prediction, MS2 level interference correction and global TIC (total intensity count) mediated normalization was carried out. 1% FDR cut-off was applied on precursor and peptide level to ensure true detection of MS signals. The extracted ion chromatogram (XIC) intensity of peptides was compiled to quantify the normalized protein abundance values which was extracted for downstream quantitative analysis. The ZenoSWATH data is available via ProteomeXchange with identifier PXD065012.

### 2.12 Affinity tag-based immuno-precipitation

The HTR8/SVneo cells were transfected with flag-tagged human LBX1 construct and then harvested in IP lysis buffer (25 mM Tris-HCl pH 7.4, 150 mM NaCl, 1 mM EDTA, 1% NP-40 and 5% glycerol) added with 1X halt-protease and phosphatase inhibitor cocktail (78441, Thermo Scientific) post 48-hour incubation. The wells transfected with empty vector were used as controls. The lysates were centrifuged to remove cell debris followed by quantification of the extracted protein using BCA protein assay kit (23225, Pierce, Thermo Scientific). 1 mg lysate from each sample were taken and incubated with 45 µl pre-equilibrated anti-Flag M2 Magnetic beads (M8823, Millipore) at 4 °C for overnight. Post incubation the magnetic beads were washed with IP lysis buffer 3 times, followed by a single wash with milli-Q water. Finally, the enriched proteins were eluted two times in 20 µl 2X lamellae dye (without β-mercapto ethanol) by heating the beads at 95 °C. The eluted proteins were next loaded onto 12 % sodium dodecyl sulphate-polyacrylamide (SDS-PAGE) gel for either western blot analysis or in-gel digestion.

### 2.13 In-gel sample preparation

The immuno-precipitated samples were resolved 30% at 100 V in an SDS-PAGE gel, followed by staining and de-staining. The gels were washed using milli-Q water to remove residual acetic acid and then the whole sample lane were excised and divided in three fractions using sterile scalpels carefully. The excised gels were diced into 1 mm^3^ pieces, washed in 50% acetonitrile and 50 mM ABC buffer for proper stain removal and then dehydrated using 100% acetonitrile. Next, the samples were hydrated using 150 µl of reduction solution (10 mM DTT in 100 mM ABC), followed by incubation at 56 °C for 45 minutes. 100 µl solution having 20 mM IAA in 100 mM ABC were added to each tube and incubated for 1 hour at room temperature for alkylation. The gels were again washed with 50% acetonitrile and 50 mM ABC buffer and dehydrated to remove the excess solutions. Then, 150 µl of 10 g/ml MS-grade trypsin (90057, Pierce, Thermo Scientific) were added to each tube and incubated at 37 °C. After overnight trypsin digestion, the peptides were eluted in an extraction buffer (60% acetonitrile and 0.1% formic acid) and the three fractions from one sample was pooled into a single tube, which was further vacuum dried. The salts from the eluted peptides were removed using Oasis HLB C18 cartridges (WAT094225, Waters), followed by vacuum-drying for −80 °C storage.

### 2.14 Data dependent acquisition and processing

The LC-MS/MS profiling of the tryptic peptides were performed using Sciex ZenoTOF 7600 mass spectrometer connected with Waters microscale LC system (ACQUITY UPLC M-Class System). An Equivalent amount of digest from each sample were initially loaded onto Luna 5 µm C18 Micro trap column (20 mm X 0.3 mm, 100 Å, Phenomenex) and finally resolved using a nanoEase M/Z HSS T3 analytical column (150 mm X 300 µm, 1.8 µm, 100 Å, Waters,) with a constant flow rate of 5 µL/min. A binary solvent manager was used to maintain an increasing linear gradient of solvent B (100% (v/v) acetonitrile with 0.1% (v/v) formic acid) till 37 minutes followed by washing and equilibration with solvent A (2% (vol/vol) acetonitrile, 0.1% (vol/vol) formic acid) for the remaining of 50 minutes. A data dependent acquisition (DDA) scheme was followed using optimized settings for voltage, curtain gas, nebulizer gas, heater gas, and the source temperature. The peptides were subjected to survey scans (MS1) (m/z 300-1500 Da) for 100 msec followed by selection of top 45 ions for subsequent MS2 (200-1800 m/z) scans each for 20 msec, adding a cycle time of 1.24 sec. A 5 sec dynamic exclusion window and a minimum 30 ppm tolerance were utilized for initial survey scans. MS1 and MS2 fragmentation were conducted using a dynamic collision energy with a spread of 5 and a declustering potential of 80.

The acquired raw DDA (.wiff) files were processed in MSConvertGUI from Proteowizard^11^ to generate the mzML files. These mzML files were loaded onto MSfragger (v4.4.1)^12^ to search the data in default LFQ-MBR workflow against the UniProtKB human protein database (December,2024) incorporated with isoforms and reversed sequences. Briefly, “stricttrypsin” enzyme specificity with maximum 2 missed cleavages were selected for subsequent modification filter where fixed modification of cysteine Carbamidomethylation (+57.0215 Da) and variable modification of methionine oxidation (+15.9949 Da, maximum 3 occurrences), and N-terminal acetylation (+42.0106 Da, maximum 1 occurrence) was used for peptides (500-5000 Da) of 7 to 50 amino acids lengths. The extracted peptide spectrum matches (PSMs) were mapped onto peptides and proteins using 1% FDR filtering criteria. Finally, the MS1-based label free quantitation was done using high confidence peptide matching proteins across the samples, with match between run (MBR) algorithm enabled. Further, to identify true interactors Significance Analysis of INTeractome (SAINT) was done using SAINTexpress^13^ plugin in Fragpipe (v23.0). The default parameters were used for probabilistic scoring that identifies the true protein-protein interactions while filtering out contaminants using background control. The exported spectral count file was used for further analysis. This AP-MS data is available via ProteomeXchange with identifier PXD075722.

### 2.15 Immuno-histochemical analysis

Paraffin-embedded tissue blocks were first sectioned into thin slices using a Leica microtome. The obtained sections were carefully transferred onto pre-coated poly-L-lysine slides to facilitate proper adhesion during subsequent staining procedures. Immunohistochemical staining was then carried out following the manufacturer’s protocol using the Ultra Vision Quanto Detection System HRP DAB kit (TL-125-QHD, Thermo Fisher Scientific). The sections were incubated with the primary anti-LBX1 (1:500, PA5-68985, Invitrogen) antibody and after completion of the staining procedure, the slides were mounted using DPX mounting medium. Representative images of the stained sections were subsequently captured using an Olympus IX300 microscope.

### 2.16 Immuno-fluorescent staining of cells

1X10^5^ cells were seeded on cover slips in a 12 well plates and incubated till the confluency reaches to 65%. Then, transfection and Importazole treatment was performed following the similar protocol discussed previously. Next, the cover slips were fixed with 4% paraformaldehyde for 10 minutes at RT and then transferred to humidified chambers for immunostaining. The immuno-staining was performed using antibody against LBX1 (1:400, PA568985, Invitrogen) and KPNB1 (1:500, MA3-070, Invitrogen) for 1.5 hours at RT, followed by 3 times PBS wash and secondary antibody (1:500, Alexa fluor-594 goat anti-mouse lgG and Alexa fluor-684 goat anti-rabbit lgG, Thermo Fisher Scientific) incubation at RT for an hour. Post PBS wash 60 μL DAPI from 5 ug/ml stock solution was added to each cover slip and then incubated for 5 minutes. After, the final PBS wash the cover slips were kept for drying and the dried cover slips were mounted in a glass slide and stored at −20 °C. The slides were imaged in a confocal microscope (Leica TCS SP8) and processed in ImageJ software to analyse the localization of LBX1 post IPZ treatment.

### 2.17 Immuno-blotting

The immuno-precipitated samples and the transfected cell lysate post IPZ treatment were subjected to western blots. Briefly, the total pull-down elutes and 30 µg of total protein samples were loaded in the individual wells of 12% SDS-PAGE gel which was electrophoretically resolved at 100 V. Next, the resolved proteins were transferred onto a 0.45 µ polyvinylidene fluoride (PVDF) membrane (IPVH00010, Millipore, Merk) followed by 1 hour of blocking with 5% (W/V) skimmed milk (GRM1254, Himedia). The respective blots were overnight incubated at 4 °C with primary antibodies against LBX1 (1:1000, PA5-68985, Invitrogen); KPNB1 (1:1000, MA3-070, Invitrogen), mTOR (1:5000, 66888-1-Ig, Proteintech); POLR1E (1:1000, 16145-1-AP, Proteintech); RPL7A (1:2000, 15340-1-AP, Proteintech); RPS6 (1:1000, 66886-1-Ig, Proteintech); FN1 (1:1000, 15613-1-AP, Proteintech); VIM (1:1000, 10366-1-AP, Proteintech); Flag (1:1000, F3165, Millipore); BiP (1:1000, 3177, CST); ATF6 (1:1000, 65880, CST); PERK (1:500, bs-2469R, Bioss); IRE1α (1:1000, 3294, CST); GAPDH (1:10000, A19056, ABclonal) and beta-actin (1:5000, 4970, CST). Next, the blots were washed with tris-buffered saline-tween (TBST) (0.1% [vol/vol]) and incubated with HRP conjugated secondary anti-rabbit or anti-mouse antibody (1:10000 & 1:10000 respectively) for an hour at room temperature. After TBST wash, the blots were developed using Immobilon Forte Western HRP Substrate (WBLUF, Millipore, Merk) and the digital image were captured using Image Quant LAS 4000 (GE Healthcare Bio-Sciences AB, Sweden). The respective band intensities were quantified using Fiji (v2.17.0) software and the relative protein expression was subsequently normalized using either GAPDH or beta-actin band intensity.

### 2.18 Bioinformatic and statistical analysis

Clinical characteristics of the enrolled study population were summarized using median (IQR) for continuous variable and percentage for categorical variable. The quality of raw data was assessed with Pearson’s coefficient, principal component analysis (PCA), partial least square discriminant analysis (PLS-DA), coefficient of variance (CV) statistics, followed by median and IQR calculation. R package Factoextra (v1.0.7) was used for PCA analysis, while PLS-DA was computed using Metaboanalyst (v6.0) webtools. All other calculations were performed using base R (v4.3.1) functions and visualized using the ggplot2 (v3.5.2) in Rstudio (v2023.09.1+494), if not either mentioned. All the pathway enrichment was performed using ClusterProfiler (v4.10.1) package using ReactomePA (v1.46.0) or gene ontology (GO) database. Receiver-operating characteristic (ROC) analysis was used to determine the predictive threshold of selected proteins from placental SWATH data and plasma HR-MRM data by utilizing the pROC (v1.18.5) and Boruta (v9.0.0) packages in R. The heatmaps and upset plots were generated by pheatmap (1.0.12) and UpsetR (1.4.0) package respectively. All the graphical illustrations were created using BioRender (https://www.biorender.com/).

The differential protein abundance in SWATH as well as in HR-MRM across the clinical groups (EPTB vs TB and LPTB vs TB for SWATH and sPTB vs TB for HR-MRM) and cellular samples (OE vs COE and KD vs CKD) was estimated using two-tailed, unpaired Student’s t-test. An absolute protein abundance of ≥ 1.5-fold with FDR adjusted p-value (q-value) of ≤ 0.05 was considered differentially expressed. For western blot data, statistical analysis was performed using Prism 8 for all the experiments. The datapoints were plotted at least for three biological replicates and expressed as mean ± Standard error of mean (SEM). The statistical significance between groups were measured using student’s t-test where P values <0.05 was considered as statistically significant and are indicated by asterisks as follows: *P<0.05, **P<0.01, ***P<0.001, ****P<0.00013.

## Results

### 3.1 Clinical and socio-demographic representation of study participants

The placenta proteomics study was conducted using 70 participants, who were selected from the GARBH-INi cohort using a set of inclusion and exclusion criteria (**Supplementary Figure S1A**). A cross-sectional study design was employed to profile individual placenta samples over a POG range, which will capture the placental aging signatures essential for distinguishing sPTB specific regulations. Recent ACOG guidelines^9^ were followed to classify 12 participants in early preterm birth group (EPTB, less than 34 weeks), 22 participants in late preterm birth group (LPTB, 35-37 weeks), and 36 participants in term birth group (TB, 37-39 weeks). The EPTB and LPTB subgroups were considered as case while the TB subgroup is treated as control. No significant difference was observed for maternal age, BMI, parity, education, or occupation. While the placental weight and gestational age at delivery is significantly less for both EPTB and LPTB subgroup. Similarly, the neonatal gender and feto-placental weight were unchanged, but the birth weight of preterm babies was significantly reduced (**Table 1**).

**Table 1:** Participant’s sociodemographic and clinical characteristics at baseline and delivery, along with neonatal parameters at birth ^a^.

| **Variable** | **Early preterm**<br>**(n=12)** | **Late preterm**<br>**(n = 22)** | **Term**<br>**(n = 36)** |
| --- | --- | --- | --- |
| ***sociodemographic and clinical characteristics of participants at baseline and delivery*** |  |  |  |
| **maternal occupation** |  |  |  |
| un-employed (n, %) | 11 (100%) | 21 (95.5%) | 32 (89%) |
| un-skilled and semi-skilled worker (n, %) | 0 (0%) | 1 (4.5%) | 3 (8%) |
| clerk, shop owner, farm owner etc. (n, %) | 0 (0%) | 0 (0%) | 1 (3%) |
| **maternal education** |  |  |  |
| illiterate–primary school (n, %) | 4 (33%) | 5 (23%) | 11 (30.5%) |
| middle–hr. sec school (n, %) | 7 (58%) | 16 (73%) | 18 (50%) |
| graduate and above (n, %) | 1 (9%) | 1 (4%) | 7 (19.5%) |
| **maternal BMI at first visit (kg/m2)** | 19.75<br>(18.17,22.58) | 20.28<br>(18.07,21.82) | 19.89<br>(17.91,22.10) |
| **parity** |  |  |  |
| 0 | 8 (67%) | 11 (50%) | 22 (61%) |
| 1 | 2 (16.5%) | 7 (32%) | 9 (25%) |
| 2 | 2 (16.5%) | 3 (14%) | 4 (11%) |
| 3 | 0 (0%) | 1 (4%) | 1 (3%) |
| **gestational age at birth (weeks)\*\*\*** | 33.85<br>(32.45,34.5) | 36.1 (35.4,36.38) | 38.4 (37.5,39.03) |
| **maternal age at delivery (years)** | 21.5 (19,23.25) | 23.5 (20.25,25) | 23 (20,25) |
| **Placental weight\*** | 398.2<br>(328.55,445.33) | 404.7<br>(371.03,472.35) | 500.35<br>(409.48,549.48) |
| ***neonatal characteristics at birth*** |  |  |  |
| **birth weight–placental weight ratio** | 4.79 (3.94,5.53) | 5.38 (4.97,5.91) | 5.67 (5.08,6.63) |
| **birth weight (kg)\*\*\*** | 1.961 (1.48,2.03) | 2.33 (2.13,2.48) | 2.7 (2.58,2.93) |
| **gender** |  |  |  |
| male (n, %) | 8 (66%) | 11 (50%) | 19 (53%) |
| female (n, %) | 4 (33%) | 11 (50%) | 17 (47%) |

<sup>a</sup> Total 70 participants were selected for proteomics study with upper panel explaining demographic and clinical parameters for mothers at baseline and delivery, while lower panel explains neonatal variables. The data is represented as median (Q1, Q3) or number (percentage), counts as n (%) for continuous and categorical variables, respectively; BMI = Body Mass Index. Statistical significance is calculated either using one-way ANOVA for continuous variables, or utilizing Chi-square test for categorical variables. \*p-value < 0.05. \*\*p-value < 0.005. \*\*\*p-value < 0.0001.

The findings from placenta proteomics study were validated in maternal plasma from 18-20 weeks of gestation. A similar pipeline was followed for selection of 53 participants from the cohort (**Supplementary Figure 1B**). 26 participants were classified in spontaneous preterm (sPTB, GA <35 weeks) subgroup, whereas the other 27 are categorized in term birth (TB, GA >39 weeks) group. The distribution of socio-demographic characteristics revealed no substantial difference between maternal age, BMI, education or occupation. About 50% of the selected mothers were nulliparous (**Table 2**).

**Table 2:** The sociodemographic and clinical features of selected participants for plasma validation study ^b^.

| **Variable** | **Preterm (n=26)** | **Term (n=27)** |
| --- | --- | --- |
| **maternal occupation** |  |  |
| un-employed (n, %) | 26 (100%) | 26 (96%) |
| un-skilled (n, %) | 0 | 1 (4%) |
| **maternal education** |  |  |
| illiterate–primary school (n, %) | 6 (23%) | 10 (37%) |
| middle–hr. sec school (n, %) | 18 (69%) | 8 (30%) |
| graduate and above (n, %) | 2 (8%) | 9 (33%) |
| **maternal BMI at first visit (kg/m2)** | 18.42(17.51,22.68) | 20.32(18.74,22.86) |
| **parity** |  |  |
| 0 | 13 (50%) | 13 (48%) |
| 1 | 10 (38%) | 10 (37%) |
| 2 | 2 (8%) | 3 (11%) |
| 3 | 0 | 0 |
| 4 | 1 (4%) | 1 (4%) |
| **gestational age at birth (weeks)\*\*\*\*** | 32.8(31.48,34.33) | 39.4(39.3,39.8) |
| **maternal age at delivery (years)** | 21.5(20,23.75) | 23(21,26) |

<sup>b</sup> We selected 53 participants from the cohort for placenta proteome validation using the targeted MS platform. The maternal variables were represented as median (Q1, Q3) or number (percentage), for continuous and categorical variables, respectively; BMI = Body Mass Index. Statistical significance is calculated either using Student's t-test for continuous variables, or utilizing Chi-square test for categorical variables. \*p-value < 0.05. \*\*p-value < 0.005. \*\*\*p-value < 0.0001.

### 3.2 Placental proteome identifies the signatures of spontaneous pre-maturity

Placental dysfunction is a hallmark for various adverse pregnancy related outcomes. Thus, we profiled placental signatures during spontaneous pre-maturity using a SWATH based DIA workflow with the selected samples from the cohort (**Figure 1A**). We identified a total of 6,989 peptides corresponding to 1,550 protein groups and 1,660 proteins from the of placental tissue samples. Additionally, our placenta proteomics data revealed a considerable overlap with other proteomics studies on placental tissue from different adverse pregnancy outcomes, leaving only 2.3% (158) unique proteins identified in this study (**Figure 1B**). Coefficient of variation (CV) distribution of different clinical sub-groups indicated an intragroup CV of more than 35% (**Figure 1C**). While the values for mean inter-quartile range (1.92±0.079) (IQR) and median (4.38±0.296) conveys low inter-sample variability (**Supplementary Figure 2A & B**). Principal component (PC) analysis showed a left to right clustering trend with increasing POG, separated at PC1 with maximum variation of 29.1% and 6.5% at PC2 (**Supplementary Figure 2C**). Further, partial least squares discriminant analysis (PLS-DA) demonstrated a clear clustering of samples in three distinct groups, with partial overlaps between EPTB-LPTB as well as LPTB-TB groups reflecting the true nature of POG overlap between the clinical sub-groups (**Figure 1D**). Therefore, the IQR and median values, together with CV statistics, PCA, and PLS-DA, indicated high quantitative precision and robust biological signal, sufficient for downstream differential analysis. Sub-cellular localization analysis using DeepLoc (v2.0) webtool^14^ enriched cytoplasmic proteins (802) as the most represented cellular location (**Supplementary Figure 2D**). Next, we calculated the cellular heterogeneity of placental tissues based on the abundance of unique cell type markers, as described earlier^15^. The unique marker proteins specific to decidual cells (DE), extravillous trophoblasts (EVT), fibroblasts (FB), Hofbauer cells (HC), syncytiotrophoblasts (SCT), villous cytotrophoblasts (VCT) and villous endothelial cells (VEC) were selected and verified from the trophoblast cell proteome data in human protein atlas and literature curated placental single-cell RNA-sequencing data^16^ (**Figure 1E & Supplementary File S1**). The analysis revealed VECs as most abundant cell type whereas the SCTs were found to be most abundant among the trophoblastic lineages (**Supplementary Figure 2E**). However, the individual cell type abundance did not show any significant changes between the EPTB, LPTB and TB groups, thus precluding cellular heterogeneity as a contributing factor for preterm phenotype (**Supplementary Figure 2F & Supplementary File S1**).

**Figure 1:**
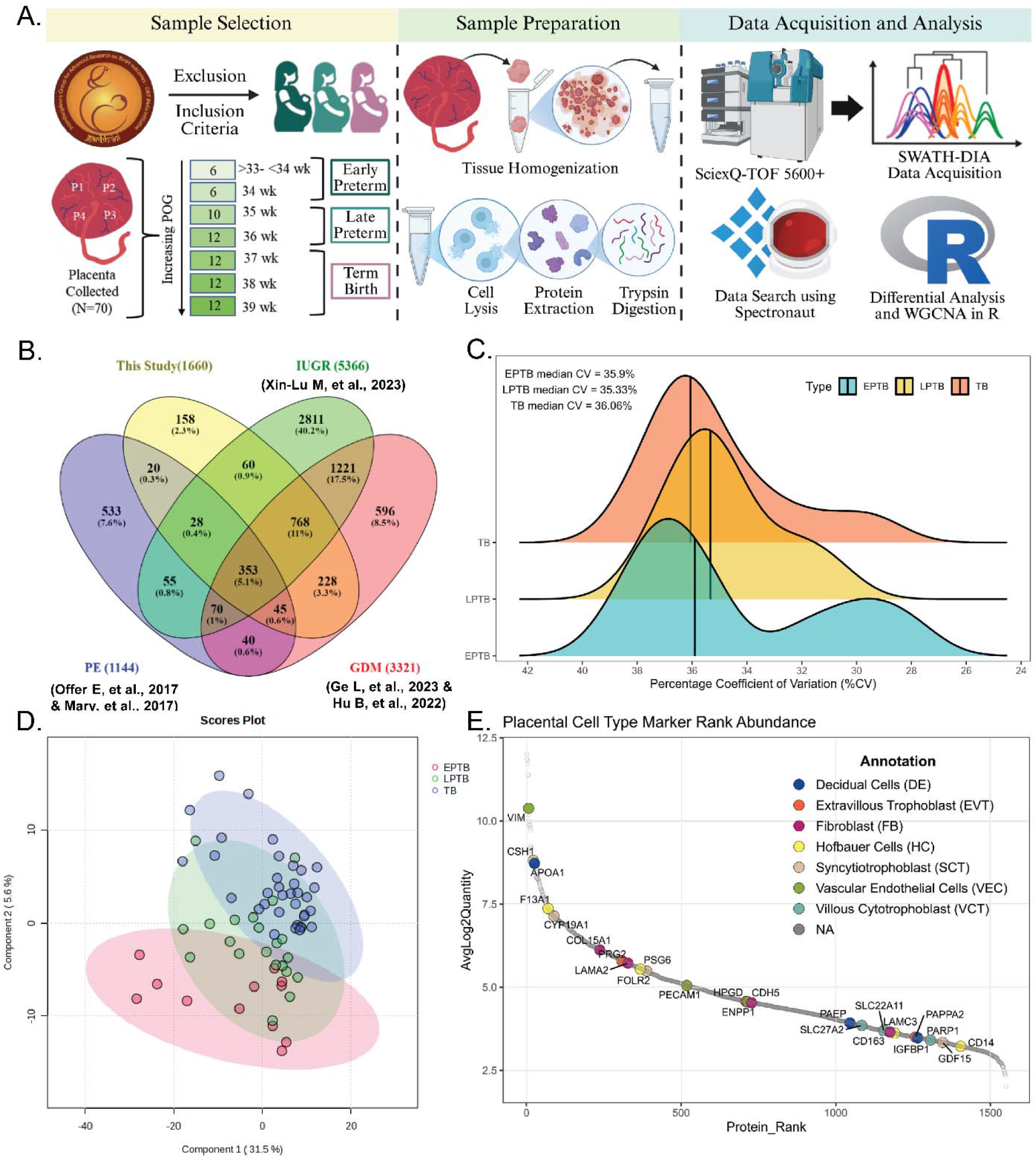
Proteome landscape of placental prematurity due to spontaneous preterm birth. **A)** The proteomic workflow followed for placental profile of 70 participants with spontaneous preterm. **B)** Venn diagram showing overlap between identified proteins of this study with other placenta proteomes from different APOs. **C)** CV distribution of EPTB, LPTB and TB subgroup samples. **D)** Clustering of different comparison groups using PLS-DA with component 1 and 2 plotted on x and y axis respectively. **E)** Rank plot showing abundance distribution of identified cell type markers in the placenta proteome.

### 3.3 Spontaneous preterm birth significantly alters the placental proteome dynamics

We next identified the differentially expressed protein (DEPs) signatures of placental prematurity utilizing a statistical threshold for q-value of <0.05, which revealed 14 up-regulated and 17 down-regulated proteins in the LPTB vs TB comparison (**Figure 2A & Supplementary File S2**). Similarly, 41 proteins were upregulated and 32 were downregulated when LPTB was compared with the TB group (**Figure 2B & Supplementary File S2**). The overlap between these regulated proteins from LPTB vs TB and EPTB vs TB comparison highlighted 11 shared proteins (**Figure 2C**), among which MSLN, EMILIN2, ACADS, FTH1, MYL1, MYLPF were downregulated and PDLIM5, CYP1A1, KRT9 were elevated in both comparisons. While ABI3BP and SAA1 showed up-regulation in LPTB and down-regulation in EPTB condition (**Figure 2D**).

**Figure 2:**
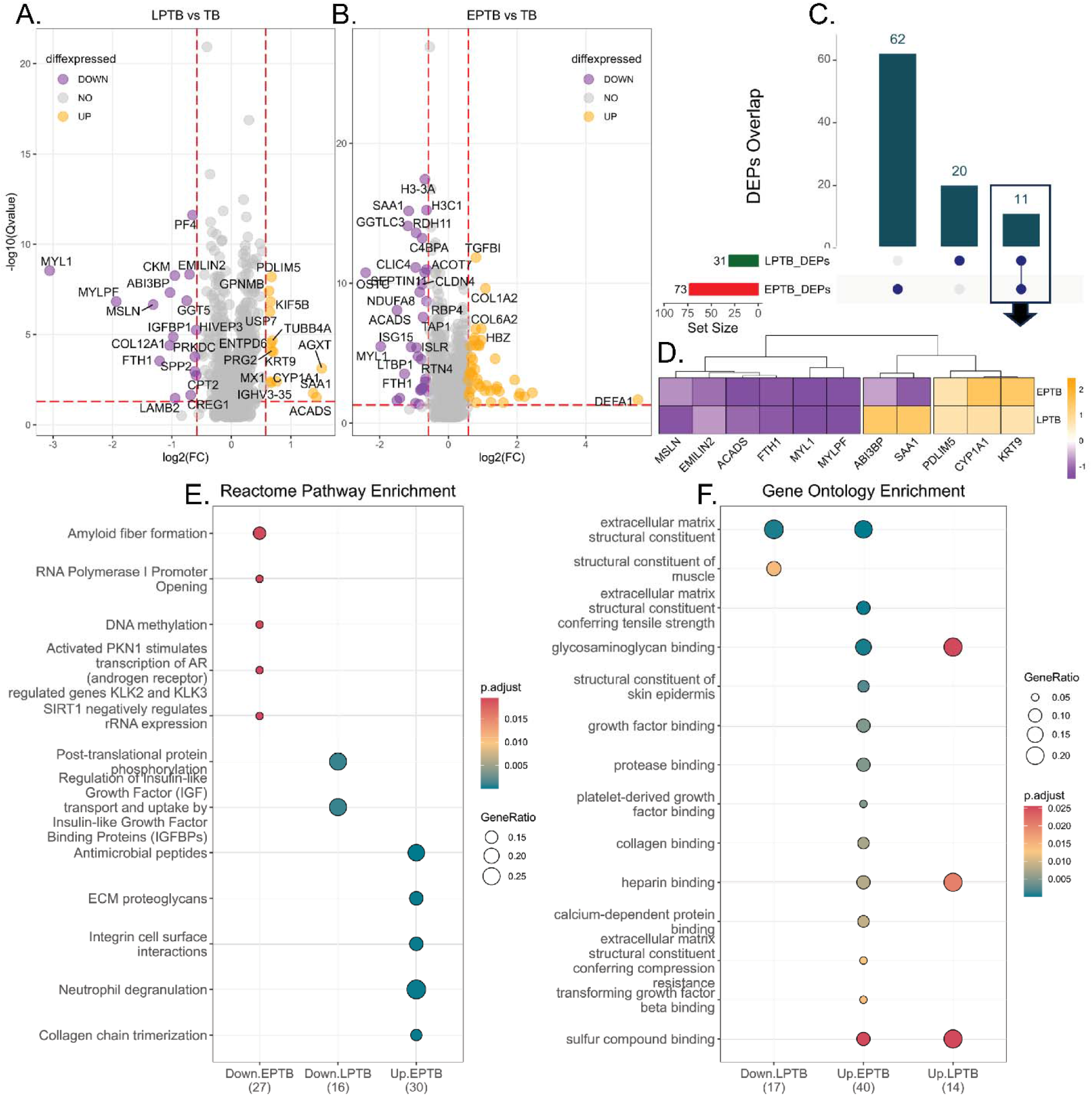
Differential protein abundance caused by spontaneous preterm, along with their functional enrichment. The volcano plot showing comparison between **A)** LPTB vs TB and **B)** EPTB vs TB subgroup, where up (orange) and down (purple) regulated proteins are identified using absolute log2 FC (x-axis) and q-value (y-axis) cut-off of >0.6 and <0.05 respectively, denoted using red-dotted lines. **C)** The upset plot shows the common and uniquely regulated proteins from the comparison of the previously mentioned subgroups. **D)** The 11 common DEPs are highlighted using heatmap with orange denoting higher z-score abundance and purple denoting lower. The pathway enrichment of all the DEPs using **E)** Reactome or **F)** GO database shows differentially regulated pathways from each comparison. The pathways are represented on y-axis with corresponding circle colour and size denoting adjusted p-value and gene ratio respectively. The x-axis shows the regulatory direction of pathways in each group with the associated number of proteins used in enrichment, mentioned in the bracket.

Reactome pathway enrichment of all the DEPs revealed the following terms: RNA polymerase I promoter opening, DNA methylation and PKNI1 stimulated transcription of androgen receptor regulated genes downregulated in EPTB; post-translational regulation of IGF and transport and uptake by IGFBPs downregulated in LPTB; ECM proteoglycans, integrin cell surface interactions, neutrophil degranulation and collagen chain trimerization upregulated in EPTB (**Figure 2E & Supplementary File S2**). Similarly, gene ontology enrichment of the DEPs revealed glycosaminoglycan binding, heparin binding and sulphur compound binding to be upregulated in both EPTB and LTPB whereas extracellular matrix structural constituent showed upregulation in EPTB and downregulation in LPTB (**Figure 2F & Supplementary File S2**).

### 3.4 Co-expression network highlights preterm associated hub proteins in placenta

The comparison between mature term placenta (GA >37 weeks) with a less mature preterm placenta (GA <37 weeks) come with a shortcoming of also capturing maturity related proteome regulation. Thus, we performed weighted gene correlation network analysis (WGCNA) to reveal the co-expression patterns of the proteins, that are expected to be in-sync with the changes in their expression trajectories over the POG. Similar strategies have been followed in other clinical tissue proteomics studies^17–20^. Therefore, we built a co-expression network using a matrix of the 1550 identified proteins and their relative abundance from 70 samples by obtaining a scale free topology with a soft thresholding power (β) of 7, selected based on scale independence (R^2^ = 0.889) and mean connectivity (Mean K = 7.16) calculation (**Supplementary Figure 3A**). Initially, the network generated 7 modules, which are subsequently regrouped based on high relevance of module eigengenes with adjacent modules, yielding a total of 5 modules for downstream analysis (**Figure 3A**). The interactive relations between the modules and their respective proteins were visualized on a heatmap based on topological overlap matrix (TOM) (**Supplementary Figure 3B**). Finally, the protein number clustered in each module were reported (**Supplementary Figure 3C**), followed by the hierarchical clustering of the modules (**Supplementary Figure 3D**).

**Figure 3:**
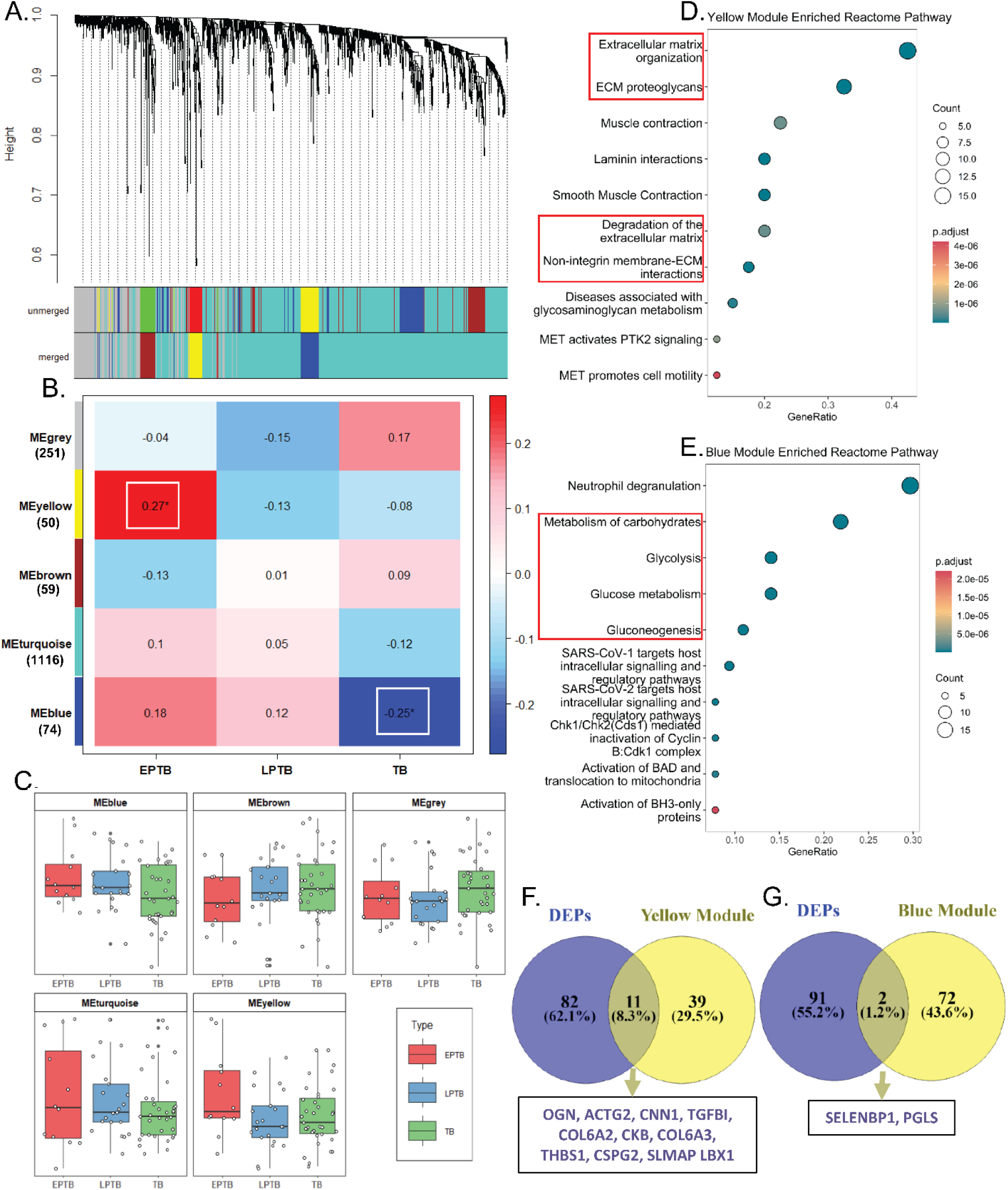
Protein co-expression network generation to identify the hub proteins. **A)** The clustered dendrogram showing the grouping of 1550 identified proteins initially in 7 different colour modules (unmerged), which is ultimately merged to 5 independent co-expression modules. **B)** The module-trait relationship is identified using co-relation matrix where positive correlation is indicated using red and negative correlation is denoted with blue. The significant correlations (p-value <0.05) are mentioned with a star. The different coloured modules with their respective protein numbers are shown in y-axis, while the clinical sub-groups are mentioned in x-axis. **C)** The calculated module eigengene (ME) of each sample for a coloured module were distributed in EPTB, LPTB and TB sub-groups and represented with a box-plot. The Reactome pathway enrichment of **D)** yellow module and **E)** blue module proteins are shown with the circle size and colour denoting the pathway associated protein count and their adjusted p-values, respectively. The Venn diagram showing overlap of DEPs with either **F)** yellow module or, **G)** blue module proteins. The common proteins are highlighted in the lower box.

We next explored the module-trait correlation between the identified modules and the clinical groups to reveal a relatively positive correlation of yellow module with EPTB condition (R^2^ = 0.27 and P<0.028) and a significant negative correlation with blue module for TB outcome (R^2^ = −0.25 and P< 0.042) (**Figure 3B**). The module eigengene (ME) values of a sample from the respective clinical groups were plotted in a boxplot for all the five identified modules to demonstrate their relative distribution (**Figure 3C**). As, only the yellow and blue module showed significant correlation with the clinical traits, we further explored the functional relevance of the proteins in this module only (**Supplementary File S3**).

To evaluate the biological functions altered in the yellow and blue protein modules, we performed Reactome pathway enrichment analysis. The yellow module proteins were significantly enriched in Reactome pathways associated with extracellular matrix (ECM) dynamics, including ECM organization, ECM proteoglycans, ECM degradation, and non-integrin membrane-ECM interactions (**Figure 3D & Supplementary File S3**). In contrast, blue module proteins were significantly enriched in pathways related to carbohydrate metabolism, including glycolysis, glucose metabolism, and gluconeogenesis (**Figure 3E & Supplementary File S3**).

We further assessed protein significance within the selected modules using scatter plots, which revealed clear upregulation trends in the yellow module and downregulation trends in the blue module. The top 10 most represented proteins were labelled in the scatter plot (**Supplementary Figure 3E & F**). Next, we merged the proteins from these two significant modules with the DEPs, identifying 13 common proteins that were elevated in samples with adverse outcomes (**Figure 3F & G**). From the yellow module, we identified 11 proteins, including mimecan (OGN), actin gamma-enteric smooth muscle (ACTG2), calponin-1 (CNN1), transforming growth factor-beta-induced protein ig-h3 (TGFBI), collagen alpha-2(VI) chain (COL6A2), collagen alpha-3(VI) chain (COL6A3), creatine kinase B-type (CKB), thrombospondin-1 (THBS1), versican core protein (VCAN), sarcolemmal membrane-associated protein (SLMAP), and transcription factor LBX1 (LBX1). Similarly, from the blue module, methanethiol oxidase (SELENBP1) and 6-phosphogluconolactonase (PGLS) overlapped with the DEPs. We then plotted the longitudinal expression trends and calculated the AUROC values for these 13 hub proteins (**Supplementary Figure 4A-J & 5A-C**). A compiled AU-ROC values for EPTB and LPTB outcome were represented in the (**Supplementary Figure 5D**), while these key hub proteins were taken to next analysis steps.

### 3.5 Placenta identified hub-proteins are also regulated in maternal plasma

Placental secretory proteins may drive systemic alterations in adverse pregnancy outcomes. These differentially expressed hub proteins likely appear in plasma early POG influencing maternal physiology. Thus, we profiled their expression in maternal plasma at 18-20 weeks of POG using a targeted MS based methodology to detect their presence and identify their regulatory (**Figure 4A**). The peptide transition intensities across the 53 sPTB and TB samples clustered both the clinical sub-groups using Pearson’s correlation coefficient calculation (**Supplementary Figure 5F**) and principal component analysis (**Supplementary Figure 5E**). The differential analysis using the before mentioned cut-off highlights 13 elevated and 7 down-regulated transitions in sPTB vs TB comparison (**Figure 4B & Supplementary File S4**). The normalized transition intensity across samples was plotted in a heatmap which separates two apparent clusters of peptide transitions based on their regulatory pattern (**Figure 4C**). The proteins, containing at-least one exclusive peptide transition showing up-regulatory trend, were selected for predictive classification to discriminate between sPTB and TB samples (**Figure 4D**). We assessed the predictability of the selected peptide transitions using both receiver operating characteristic (ROC) analysis and Boruta feature selection. Predictability was quantified by the area under the ROC curve (AUROC), with features selected based on their importance scores from random forest modelling (**Figure 4E & F**). Our analysis identified THBS1, TGFBI, and LBX1 as the top predictors that distinguish sPTB from TB samples. Interestingly, we found consistent upregulation of LBX1, a transcription factor predominantly expressed in muscle in both placental tissue and plasma samples from spontaneous preterm deliveries. LBX1 effectively classified preterm cases from term controls in both sample types.

**Figure 4:**
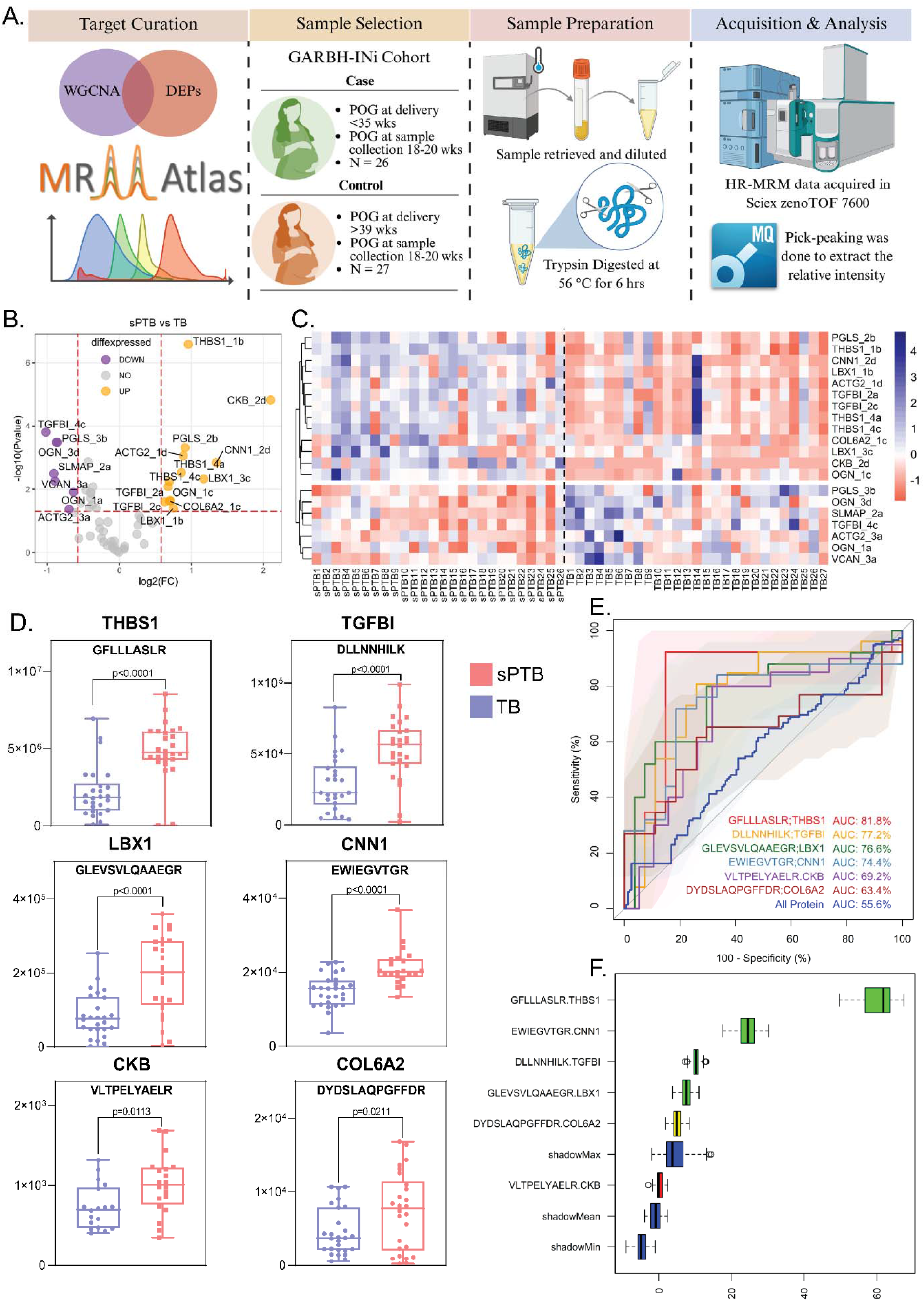
The quantification of placenta identified hub-proteins in maternal plasma. **A)** The workflow followed for identification and quantitation of the selected proteins in mother’s plasma samples from gestational age of 18-20 weeks. **B)** The volcano showing differential transitions of sPTB vs TB comparison. The x-axis and y-axis represent log2 FC and -log10 p-value with their respective cut-offs highlighted with red dotted lines. **C)** The scaled abundance distribution of regulated transitions among the sPTB and TB samples are visualized using a heatmap. The blue and red colour denotes high and low values of scaled-abundance respectively. **D)** The box-plots shows the abundance of selected peptide transitions of proteins filtered using specific criteria and their significance between the comparison groups (sPTB vs TB). The predictive tolerance of selected transitions is calculated using **E)** AU-ROC and **F)** Boruta. The x and y-axis of AU-ROC plot denotes specificity and sensitivity respectively, while the x-axis of Boruta plot shows the distribution of feature importance score.

### 3.6 LBX1 associated proteome shift in trophoblast cells reveal an alteration of key signals

Ladybird homeobox 1 (LBX1) is a transcription factor found abundantly in embryonic stem cells^21^, muscles^22^ and neurons^23^. It plays important role in embryonic development by regulating cellular differentiation^24^, migration^22^ and energy metabolism^25^. However, the presence and localization of LBX1 in placental tissues are still unclear. Therefore, we performed immuno-histochemical staining of placenta from both term and pre-term delivered mothers, to reveal elevated abundance of LBX1 in trophoblast cells of preterm placenta (**Figure 5A**). Further, the western blot analysis confirms its expression in human extra-villus trophoblast cells (HTR8/SVneo), murine placenta, human placenta and in mouse myoblast (C2C12) cells, which was used as positive control (**Figure 5B**).

**Figure 5:**
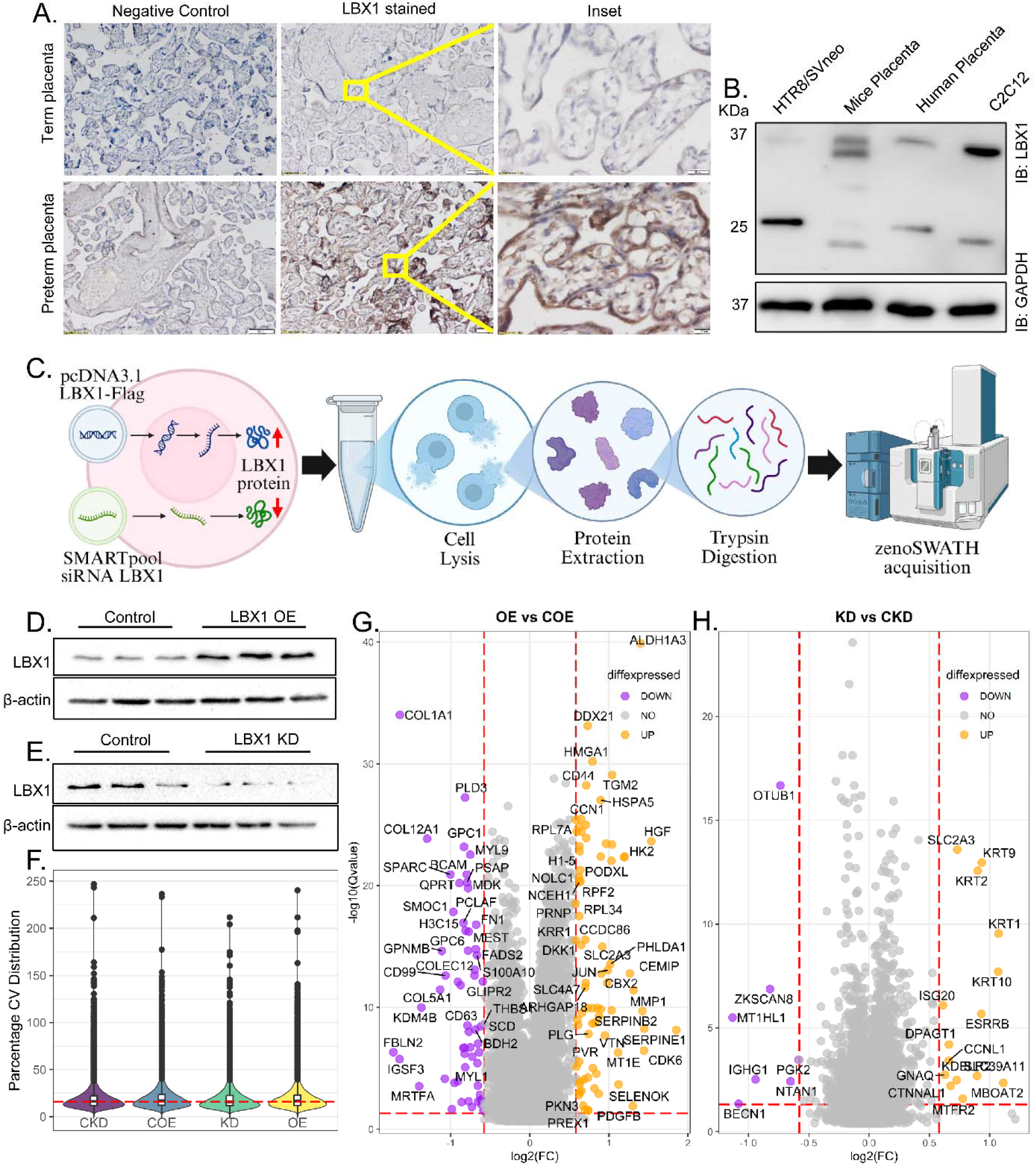
The expression of LBX1 and its regulation induced dynamic proteome in placental trophoblast cells. **A)** Representative images from immune histochemical analysis of placental slides from sPTB (lower panel) and TB (upper panel) sub-groups. The left panel shows negative staining, while the middle and right panel demonstrates LBX1 staining of placental trophoblast layers in 10X and 60X magnification respectively. **B)** The western blot analysis confirms the expression of LBX1 in HTR8/SVneo, mice placenta, human placenta and C2C12 samples. GAPDH is shown for equal loading. **C)** The schematic pipeline showing the workflow for proteomic identification of LBX1 functionality in extra-villous trophoblast (HTR8/SVneo) cells. The **D)** over-expression and **E)** knock-down of LBX1 is confirmed using immuno-blot. Here, β-actin is used as loading control. **F)** The percentage CV distribution of knock down (CKD, KD) and over-expression (COE, OE) samples post MS acquisition. The volcano plot shows differential expression of proteins in **G)** OE vs COE and **H)** KD vs CKD comparison groups, while the red dotted lines highlight the cut-off for log2 FC (x-axis) and -log10 q-values (y-axis) respectively.

Next, we explored the functional role of LBX1 in trophoblasts using HTR8/SVneo cells as a model cell line. We performed a transient over-expression as well as knock-down of LBX1 in EVTs followed by differential proteomics analysis to capture the regulatory role of LBX1 in trophoblast homeostasis (**Figure 5C**). LBX1 over-expression and knockdown were confirmed using western blot analysis (**Figure 5D & E**). The samples confirmed by Western blot were further analyzed using quantitative proteomics in ZenoSWATH mode. The SWATH analysis identified 45,875 proteolytic peptides mapping to 5140 protein groups in the spectral library of extra-villus trophoblast cells. We identified 5071 in OE and 5095 protein groups in KD conditions. Of these, 4954 proteins were common to both conditions, while 117 and 141 proteins were unique to OE and KD, respectively (**Supplementary Figure 6B**). The PC analysis separated over-expression (OE) and its control (COE) samples with PC1 and PC2 variance of 21.11% and 14.19% respectively. However, it failed to segregate the knock-down (KD) and its respective control (CKD) samples (**Supplementary Figure 6A**). Subsequent CV statistics also revealed a low median CV percentage for CKD (16.2%), COE (16.9%), KD (15.9%) and OE (16.7%) groups (**Figure 5F**). We next performed differential analysis between OE vs COE and KD vs CKD condition to identify significant DEPs using absolute fold change and q-value cut-off of ≥1.5 and ≤0.05 respectively. The relative assessment between OE and COE groups yielded 77 up-regulated and 58 down-regulated features (**Figure 5G & Supplementary File S5**). However, the KD vs CKD comparison highlighted only 22 (up-regulated 15 and down-regulated 7) regulated proteins (**Figure 5H & Supplementary File S5**). The overlap between the DEPs from both the comparison groups revealed only 4 shared features with similar regulatory direction except for KDELR2 protein (**Supplementary Figure 6C**).

Regulated proteins in LBX1 knockdown and overexpression conditions showed coordinated changes in signalling cascades. To explore the complex signalling nodes, we created a clustered expression map using z-score abundances of differentially expressed proteins from OE vs. COE and KD vs. CKD comparisons. The clustered heatmap revealed six distinct regulatory clusters, each exhibiting unique expression patterns across conditions (**Figure 6A**). This analysis enriched two largest clusters such as cluster 2 (C2=45) and cluster 3 (C3=44), which showed comparable protein abundance in KD samples but highlights opposite regulatory trend between OE and COE condition. Cluster 2 comprised downregulated proteins exclusively in OE samples and enriched pathways related to cell-substrate adhesion and cellular migration (**Figure 6D & Supplementary File S6**). We validated these findings in western blots by showing significant down-regulation of fibronectin 1 (FN1) and vimentin (VIM) only in LBX1 over-expressing cells (**Supplementary Figure 7A-C**). Cluster 1 (C1=26) showed the opposite expression trend to cluster 2, with proteins upregulated exclusively in OE samples (**Supplementary Figure 6D**). This cluster enriched pathways related to coagulation, hemostasis, and wound healing (**Supplementary Figure 6E & Supplementary File S6**). Cluster 5 (C5=11) represented a distinct enhancement of protein abundance in KD group, which also significantly enriched the pathways for development and differentiation (**Figure 6C & Supplementary File S6**). However, cluster 4 and 6 did not enriched any pathways, even after compiling 13 proteins in cluster 4 (**Supplementary Figure 6F**) and 5 proteins in cluster 6 (**Supplementary Figure 6G**). Finally, we explored the cluster 3 proteins which showed a consistent elevation of abundance in OE samples and the enriched pathways involved in ribosome biogenesis, rRNA processing, rRNA metabolic process, cytoplasmic translation, ribosomal large subunit assembly and ribosome assembly. This suggests increased ribosomal protein expression due to LBX1 overexpression (**Figure 6B & Supplementary File S6**) and it was more evident when we checked the regulation of large and small ribosomal subunit proteins identified in the global proteome data. The proteins of both the ribosomal subunits revealed a higher expression in OE condition which either become comparable to control or get reversed, in case of KD (**Supplementary Figure 7D & E**).

**Figure 6:**
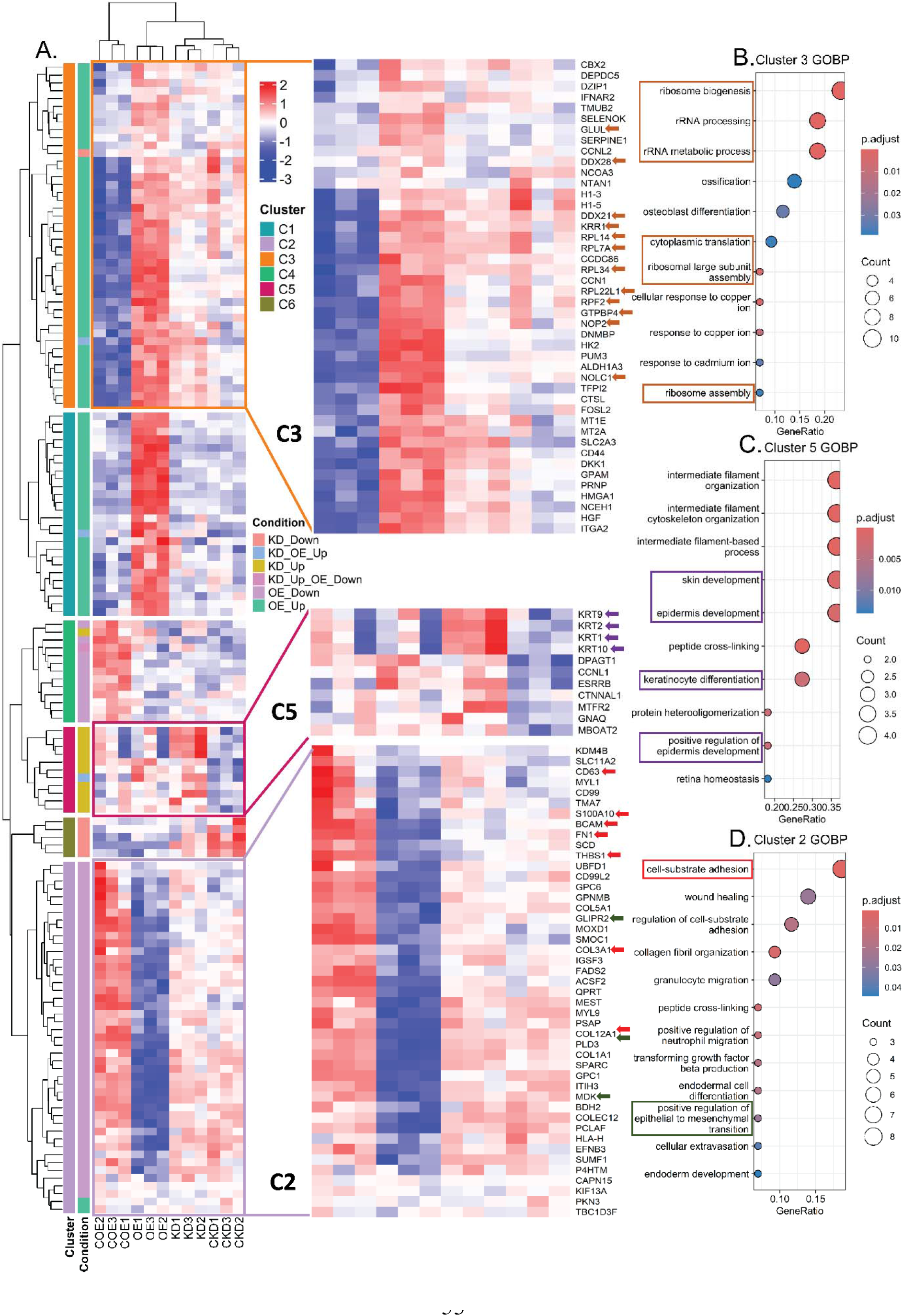
The functional enrichment of LBX1 regulated proteome. **A)** The differentially expressed proteins from OE vs COE and KD vs CKD comparisons are visualized using a heatmap plotted with z-score intensity of respective DEPs, which were further grouped by k-mean clustering. The clusters are indicated with specific colours and some (C3, C5 and C2) are further highlighted maintaining the colour schemes. The higher and lower z-scores values are denoted with red and blue colour respectively. The GO pathway enrichment of proteins from **B)** cluster 3, **C)** cluster 5 and **D)** cluster 2 shows the over-represented pathways with circle size and colour denoting associated protein count and adjusted p-values of respective pathways. The highlighted pathways and their associated proteins are indicated on the zoomed heatmap using specific-coloured arrows.

### 3.7 LBX1 regulates ribosomal functions, that elevate nascent protein synthesis and trigger unfolded protein response in trophoblast cells

LBX1 overexpression modulates ribosomal protein abundance in both subunits. However, the underlying regulatory mechanism requires further investigation. Thus, we screened the cellular interacting partners of LBX1 by flag-immunoprecipitation followed by mass-spectrometric identification of the proteins. The interactome data distinctly clustered the LBX1 IP samples from the control run using the calculated Pearson’s correlation coefficient between the samples (**Supplementary Figure 7F**). The LBX1 IP enriched proteins were scored using SAINT probabilistic scoring for background subtraction utilizing the control runs. We used the spectral count file to identify 32 true interactors using SAINT score and FC cutoff of >0.1 and 2, respectively (**Figure 7A & Supplementary File S7**). These interacting proteins revealed multiple inter-protein interactions, as identified using the STRING database (**Figure 7B**). Further, pathway enrichment of these true interacting proteins highlighted ribosome associated pathways like cytoplasmic translation and ribosomal small subunit biogenesis (**Supplementary Figure 7G & Supplementary File S7**). Importin subunit beta-1 (KPNB1) was one of the interactors which scored a high SAINT value and significance for LBX1 interaction. This observation prompted further validation using immunoprecipitation (IP) followed by Western blot, confirming a regulatory interaction between KPNB1 and LBX1 (**Figure 7C**).

**Figure 7:**
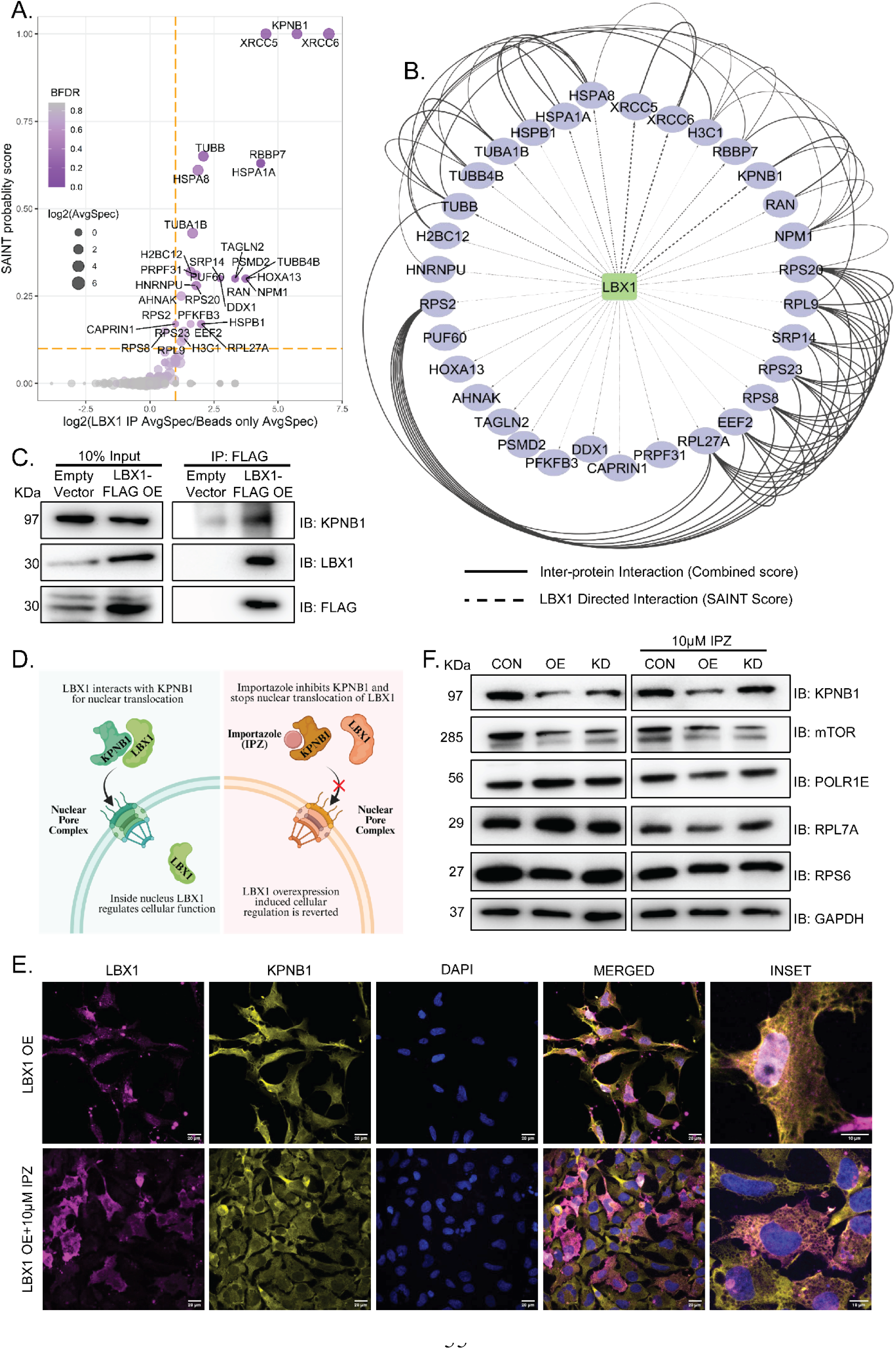
LBX1 interacts with KPNB1 to performs its ribosomal regulatory functions: **A)** The distribution of SAINT probability score and Log2 FC of average spectral counts for the identified proteins from LBX1 pull down is plotted on y and x axis respectively. The dot colour and size represent corresponding Bonferroni false discovery rate (BFDR) and log2 transformed average spectral counts. **B)** The LBX1 directed (dotted line) and STRING identified (solid line) connections between the identified proteins isolated as true interactors using SAINT scoring cut-off. **C)** Immuno-precipitation of LBX1 is performed, followed by immunoblot of enriched fractions against KPNB1, LBX1 and FLAG antibody. The empty vector is used as negative control and 50 µg protein is loaded as input to confirm the presence of target molecules. **D)** The graphical representation of KNB1 function and the mode of action for its inhibitor, Importazole. **E)** The distribution of LBX1 protein in overexpressed cells post 10 µM Importazole treatment is visualized using confocal microscopy. The cells are immune-stained using anti-LBX1 (magenta) and anti-KPNB1 (yellow) antibody, followed by DAPI staining for nucleus (blue). Images were captured using a Leica SP8 confocal microscope with a 63X oil objective. Scale bars = 20 μm. **F)** The western blot analysis of KPNB1, ribosomal proteins (RPL7A, RPS6) and ribosome regulatory proteins (mTOR, POLR1E) using LBX1 overexpression and knockdown lysate along with 10 µM Importazole treatment separately in three independent replicates. GAPDH is used as control for equal loading

KPNB1 is known to facilitate the transfer of various transcription factors from cytosol to nucleus.^26,27^ Being a transcription factor, LBX1 is also expected to be trans-located to nucleus with the help of KPNB1. Therefore, we used Importazole (IPZ), a KPNB1 inhibitor, to block its nuclear import activity.^28^ Thus, LBX1 cannot be transported to nucleus, which would diminish its ribosome associated functions (**Figure 7D**). The immuno-fluorescent assay confirms that IPZ treatment in LBX1 over-expressed cells restricts the LBX1 localization to cytosol only (**Figure 7E**). We, then carefully chose both ribosome-associated proteins (mTOR, POLR1E) and ribosomal proteins (RPS6, RPL7A) that regulate ribosome homeostasis without contributing to ribosomal structure. These targets may help to elucidate how LBX1 maintains ribosomal stability. Western blot analysis showed that LBX1 overexpression decreased KPNB1 and mTOR abundance, but IPZ treatment did not restore it. However, POLR1E and RPL7A showed elevated expression in LBX1 OE conditions, which was reversed by IPZ treatment. Finally, RPS6 showed downregulation in OE conditions, which was reversed by IPZ treatment (**Figure 7F**). The quantification of the blots showed similar observation with statistical significance (**Supplementary Figure 8A-E**).

The findings established that elevated LBX1 in trophoblast cells regulates ribosomal function. However, the ribosomal dysregulation would also affect the protein synthesis machinery in the trophoblast cells.^29^ Therefore, we probed the newly synthesized proteins in the LBX1 over-expressed cells using puromycin incorporation assay which revealed that elevated LBX1 also triggers increased synthesis of nascent proteins in the EVTs (**Figure 8A**). This nascent protein accumulation inside the trophoblast cells can induce unfolded protein response (UPR). UPR is a well-studied mechanism which is associated with various adverse pregnancy outcomes.^30–32^ Therefore, we also checked the regulation of UPR associated proteins in LBX1 over expressing trophoblast cells, which revealed higher expression of BiP and ATF6 in cells with elevated LBX1 expression. However, the abundance of IRE1α and PERK remains unchanged. The quantification and statistical analysis of the immuno-blots also report identical observations (**Figure 8B and C**).

**Figure 8:**
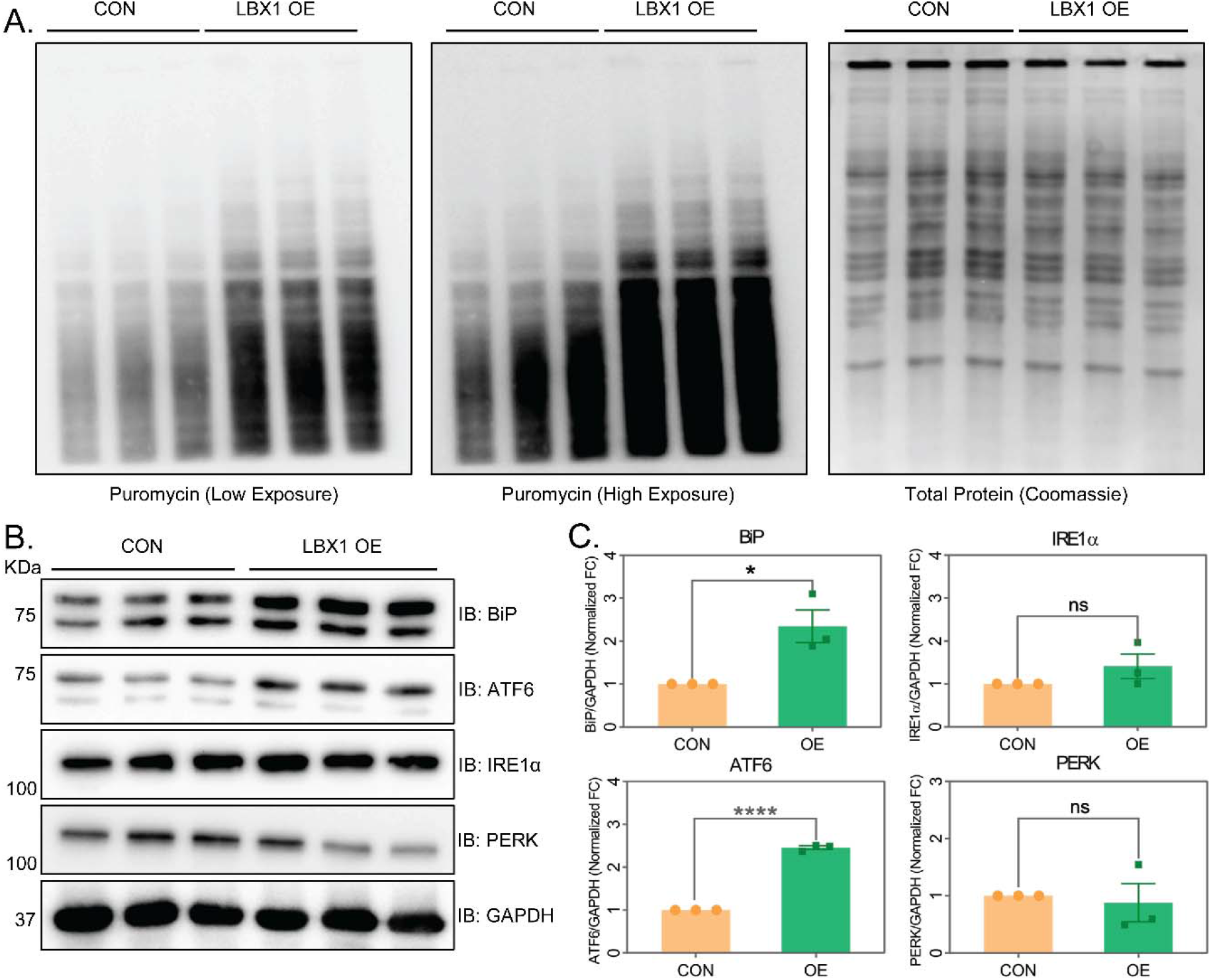
Elevated LBX1 caused increase in nascent protein synthesis triggering unfolded protein response in extra-villous trophoblast cells. **A)** The puromycin incorporation assay was performed post LBX1 overexpression using 5 μg/ml puromycin for 30 minutes and then the proteins are harvested for western blotting using anti-puromycin antibody. The total protein is visualized using Coomassie stained SDS-PAGE gel for loading control. B) The western blot analysis of unfolded protein response (UPR) related markers (BiP, ATF6, IRE1α, and PERK) are performed using three biological replicates of control and LBX1 over-expressed cells lysate. C) The densitometric quantification of the protein intensity is plotted as mean ± SEM (N=3) and the statistical significance (p-value <0.05) is calculated using Student’s t-test.

## Discussion

Preterm deliveries remain a significant challenge to the health and well-being of both the mother and child. For all the babies born premature, the underlying causes remain elusive for more than 70% of the cases and are deemed to be spontaneous^33^. There are various socio-economic, biological and genetic risk factors that can influence spontaneous preterm birth (sPTB). However, the changes in maternal physiology during pregnancy will also play a critical role in sPTB outcomes. The placenta helps maintain feto-maternal homeostasis, and its dysfunction can lead to adverse pregnancy outcomes (APOs), including sPTB^7^. Previously, studies have characterized placental metabolites^34^, DNA methylation patterns^35–37^, proteome dynamics^38,39^ and transcriptomic landscape^40,41^ to better understand the pathophysiology of sPTB. However, this literature lacks a comparative analysis between less mature sPTB placentae (GA <37 weeks) and fully mature term placentae (GA >37 weeks), thereby overlooking dynamic proteome regulation associated with placental ageing. Although a few studies have attempted gestational age correction using placental maturation signatures curated from human plasma samples^40,42^ or placental samples from non-human primates^43^ across pregnancy. However, we followed a different approach where we utilized a cross-sectional study design to track the placental aging signatures to define and distinguish the trajectory of normal placental ageing from sPTB. Therefore, we selected 70 placental samples that were collected at different gestational time points from independent pregnancies within the same cohort to characterise placental molecular signatures as a continuous function of gestational age.

The proteomic analysis identified 1,550 placental proteins, with substantial overlap with previously published datasets. This dataset showed inter-sample variability within accepted statistical limits, allowing precise quantification of biological signatures. We then stratified participants into EPTB, LPTB, and TB groups based on the gestational period at delivery to dissect sPTB-associated differences in protein expression. As the abundances of both trophoblastic and non-trophoblastic cell types were comparable across the three groups, cell-type heterogeneity is unlikely to be a major contributing factor. In contrast, differential expression analysis revealed modulation of 104 proteins in the EPTB vs TB and LPTB vs TB comparisons, with enrichment of pathways implicated in pregnancy complications, including ECM proteoglycans, neutrophil degranulation, protein phosphorylation, and integrin-mediated cell-surface interactions. Consistent findings from Gene Ontology analysis highlighted functions such as ECM structural constituents, growth factor binding, and sulfur compound binding. Differential expression analysis usually focuses on the identification of regulated proteins over certain statistical cut-offs, relying on the distribution of samples into groups, like case and control. Such a distribution often results in an unintended loss of other context-dependent information. Thus, for this cross-sectional study design we conducted WGCN analysis on the protein expression matrix. The WGCNA algorithm grouped the identified proteins into different colour modules based on their comparative expression values throughout the gestational range. The previously identified 1550 proteins were grouped in five colour modules, of which blue showed close association with the turquoise, brown associated with yellow, while the grey module, constituting unrelated proteins, did not show any associations in hierarchical clustering. Individual modules were next analysed for their association with EPTB, LPTB and TB comparison groups based on correlation coefficient, which yielded a significant positive correlation between the yellow module and EPTB, while demonstrating a significant negative correlation between the blue module and TB. This suggests that the proteins constituting the yellow module had higher expression in EPTB while those in the blue module had lower expression in TB samples. The proteins in yellow and blue modules enriched ECM organisation, degradation-related pathway and metabolism-associated pathways, respectively, all of which are important for placental homeostasis. Interestingly, when the yellow and blue module proteins were individually overlapped with the DEPs, we found 13 common proteins having a high module membership value, highlighting their robust regulation between the comparison groups. These 13 overlapping proteins showed a distinct longitudinal expression pattern over the POG that was independent of placental ageing. Interestingly, we also observed that the mean AU-ROC values of these proteins from the EPTB (mean AU-ROC, 70.3) group were higher than that of LPTB (mean AU-ROC, 57), indicating a stronger tendency to predict the early preterm outcomes.

The placental secretory factors contribute to the pathophysiology of various APOs^44^ and have the potential to be used as a discriminatory feature that can classify sPTB cases from the control population. Therefore, we performed targeted identification and quantification of these 13 key placental proteins in 53 maternal plasma samples collected at 18-20 weeks of pregnancy, to reveal the up-regulatory trends of 5 proteins, substantiating a similar observation from the placenta proteomics study. Transforming growth factor beta-induced (TGFBI) protein is one of these 5 proteins which plays a key role in cellular adhesion and migration dynamics and has been linked to gestational diabetes mellitus and sPTB^45–47^. Calponin 1 (CNN1), a thin filament-associated protein that modulates smooth muscle contraction. Its higher expression is also associated with preterm labour^48^. The other proteins are Creatine kinase B (CKB) that catalyses the reversible transfer of phosphate group between different phosphagens, such as creatine phosphate. Studies suggest that tight regulation of this enzyme is essential for a healthy pregnancy ^49, 50^. Finally, we identified Thrombospondin 1 (THBS1), which mediates cell-to-cell and cell-to-matrix interactions ^51^. Clinical studies suggest that median TBHS1 levels were higher in cervicovaginal fluid where birth occurred before term^52^. Interestingly, we found LBX1, a transcription factor needed for the development of limb muscles, diaphragm and hypoglossal cord^22^, can distinguish sPTB group from control samples due to its higher expression in both placenta and maternal plasma samples. However, its expression and localization in placental tissue have not yet been reported. Therefore, we performed immuno-histochemistry and western blots to show its presence in trophoblast layers of human and murine placentae. Further, to reveal the regulatory role of LBX1 in the placenta, we transiently over-expressed and knocked down the LBX1 proteins and performed a comprehensive quantitative proteomics study.

The extravillous trophoblast proteome showed marked changes upon LBX1 overexpression, whereas LBX1 depletion had minimal impact on proteome dynamics. This suggests that LBX1 is largely redundant under basal conditions but can drive a distinct phenotype when overexpressed. To further decode its functional role in trophoblast, we clustered the regulated proteins and performed cluster-specific pathway over-representation analysis, which demonstrates the enrichment of previously reported LBX1-associated pathways, such as cell substrate adhesion, cellular migration^22^, positive regulation of EMT^53^, skin, epidermis development, keratinocyte differentiation^24^ and coagulation pathways. We also validated the downregulation of EMT markers, FN1 and VIM due to LBX1 OE. Interestingly, we observed that LBX1 overexpression can induce upregulation of ribosome-associated proteins and their regulatory pathways. Ribosome biogenesis^54^ and the regulation of ribosomal RNA expression^55^ is linked with pregnancy complications, which influenced us to explore the ribosomal role of LBX1 in trophoblast cells. We deciphered the LBX1 mediated ribosomal homeostasis regulation by identifying 32 high confidence protein interactors of LBX1 in trophoblast cells. This highlighted the interaction of LBX1 with KPNB1, which is known to facilitate the nuclear transport of transcription factors. Thus, we used KPNB1 inhibitor, Importazole, to disrupt the nuclear localization of LBX1 which stabilizes the expression of POLR1RE, RPL7A and RPS6, that will ultimately reverse the altered ribosome biogenesis phenotype. Both POLR1E^56^ and RPL7A plays critical role in rRNA synthesis and maturation, while RPS6 facilitates the small subunit assembly^57^. mTOR plays a critical role in ribosomal homeostasis and is involved in ribosomal protein and RNA synthesis, maturation and assembly processes^58^. LBX1 is known to regulate mTOR signaling^59^ which is also evident in our western data, but IPZ treatment did not reverse its expression which indicates that mTOR is indirectly regulated by LBX1 through complex signalling cascades. Further investigation showed that LBX1 overexpression can induce nascent protein synthesis which ultimately triggers UPR in trophoblast cells. We found upregulation of molecular chaperon BiP and its one of the downstream targets ATF6 post LBX1 over-expression, which suggest that the nascent unfolded proteins are sensed by the chaperon BiP activating the downstream ATF6 pathway. The LBX1 induced UPR activation can further lead to adverse pregnancy outcome, as suggested by previous literatures.^30–32^ These observations provided a functional overview revealing a regulatory role of LBX1 in ribosomal homeostasis and its downstream activation of UPR pathways, although more experimental evidences in future will strengthen this mechanistic observation.

Finally, this study provides a comprehensive landscape of placental proteome dynamics during spontaneous preterm birth and identifies regulatory hub proteins by utilizing protein co-expression networks. This hub proteins demonstrates a considerable regulation in maternal plasma quantified using targeted MS, which ultimately highlights the presence of LBX1 in placental trophoblast cells. This directed us to explore its non-canonical ribosomal function in EVTs employing differential proteome and interactome study. The findings are validated using IPZ treatment and western blotting which demonstrates that LBX1 interacts with KPNB1 for its nuclear translocation where it regulates the abundance of key regulatory ribosomal proteins such as POLR1E, RPL7A and RPS6. This dysregulation of ribosomal homeostasis triggers accumulation of nascent proteins which activates the downstream UPR pathways leading to adverse pregnancy outcome. However, the mechanistic understanding of how LBX1 is regulating the abundance of ribosome or its associated proteins are still missing and future experiments addressing these questions will expand the understanding of non-canonical ribosomal functions of LBX1 in trophoblast cells. Moreover, how the elevated placental expression of LBX1 and its subsequent rise in maternal plasma contributes to spontaneous pre-term pathophysiology, needs more exploration. The source of LBX1 in maternal circulation and its effect on maternal physiology during pregnancy also needed further investigation. Finally, the direct or indirect factors responsible for transcriptional or translational up-regulation of LBX1 in placenta or maternal plasma needs to be identified to highlight a more detailed network of LBX1 regulation during spontaneous pre-maturity. Therefore, further investigation using murine models or clinical samples can reveal these key aspects of the current study. Despite of these limitations, this study highlights placental LBX1 as a key factor in spontaneous preterm pathology which regulates ribosomal homeostasis in trophoblast cells. In future, this LBX1 can also be used as a discriminatory feature to confidently classify spontaneous preterm from other adverse pregnancy outcomes.

## Supporting information

LBX1_Supporting Information

## Acknowledgements

We sincerely thank the research physicians, study nurses, lab technicians, field workers, internal quality improvement team, project members, and data management staff for their dedication and contributions. Special appreciation goes to the RCB mass spectrometry facility for its ongoing support. GARBH-Ini program (Interdisciplinary Group for Advanced Research on Birth Outcomes) is an initiative by the Department of Biotechnology (DBT), Government of India, coordinated by THSTI. The program brings together multidisciplinary expertise from leading research institutions like Translation Health Science and Technology Institute (THSTI), the National Institute of Biomedical Genomics (NIBMG), and the Regional Centre for Biotechnology (RCB), in collaboration with healthcare centers, including Gurugram Civil Hospital (GCH) and major New Delhi hospitals like Safdarjung Hospital and Maulana Azad Medical College (MAMC). Detailed information is available at https://www.garbhinicohort.in. We acknowledge the invaluable contribution and expertise of the DBT-funded BRIC-THSTI Biorepository in archiving and providing the biospecimens, without which this study would not have been possible.

## Author Contributions

A.B., N.K. and S.S. contributed to mass spectrometric sample preparation and data acquisition for clinical proteomics. K.S.B and A.B. did all the cell culture experiments. P.T. and M.S. performed IHC of placental sections. A.B. carried out sample preparation, data acquisition and data analysis of cellular proteomics and interactomics. N.W. and R.T. participated in clinical study monitoring and data management. D.M.S. assisted with study design. S.B. set up the cohort and provided mentorship for the clinical analyses. P.K. contributed to the implementation of the placental collection protocols, co-ordination of the Placental Program and development of study specific objectives investigating the role of placenta in sPTB within the GARBH-Ini program. T.K.M. designed and supervised overall study including proteomics, analytical methods and biochemical and proteomics data analyses. The manuscript was written by A.B. and A.T. T.K.M. and P.K. critically edited and revised the manuscript. All authors have read and approved the final manuscript.

## GARBH-Ini study group’s contribution

Shinjini Bhatnagar (S.B.), Nitya Wadhwa (N.W.), Uma Chandra Mouli Natchu (U.C.M.N.), Ramachandran Thiruvengadam (R.T.), Sumit Misra (S.M.), Dharmendra Sharma (D.S.), Kanika Sachdeva (K.S.), Amanpreet Singh (A.S.), Satyajit Rath (S.R.), and Vineeta Bal(V.B.) Alka Sharma (A.S.), Sunita Sharma(S.S.), Umesh Mehta (U.M.), and Brahmdeep Sindhu (B.S.) Pratima Mittal (P.M.), Rekha Bharti (R.B.), Harish Chellani (H.C.), Rani Gera (R.G.), Jyotsna Suri(J.S.), Pradeep Debata(P.D.), and Sugandha Arya(S.A.), Nikhil Tandon (N.T.), Yashdeep Gupta (Y.G), Alpesh Goyal (A.G.), Smriti Hari(S.H.), Aparna Sharma K (A.S.K.), Anubhuti Rana (A.R.), Rakesh Gupta (R.G.) Siddarth Ramji (S.R.) and Anju Garg (A.G.), Ashok Khurana (A.K.), Reva Tripathi (R.T.), Himanshu Sinha (H.S.), and Raghunathan Rengasamy (R.R) contributed to the clinical study design, data, and biospecimen collection, and quality assurance. Tushar K Maiti (T.K.M), Arindam Maitra (A.M.), Bhabatosh Das (B.D.), Pallavi S Kshetrapal (P.S.K.), Shailaja Sopory (S.S.), Balakrish G Nair (G.B.N), Dinakar M Salunke (D.M.S), and Partha P Majumder (P.P.M) contributed to the design of the study, supervision of laboratory experiments, and biochemical analysis.

## Data availability

The mass spectrometric data generated have been deposited in the ProteomeXchange database under accession code PXD065403 (placenta DIA data), PXD065014 (plasma HR-MRM data), PXD065012 (cell line ZenoSWATH data) and PXD075722 (LBX1 interactomics data).

## Ethics approval

The study was approved by the Institutional Ethics Committee of all participating Institutions, Translational Health Science and Technology Institute, and the Regional Centre for Biotechnology. Human Ethics Clearance, IEC protocol No: RCB-IEC-H-23. We confirm that written informed consent was obtained from all participants prior to sample collection. All bio specimens and associated clinical data were de-identified and assigned unique alphanumeric codes at the point of collection, and only coded identifiers were used during sample processing and data analysis, with the key linking codes to participant identities held securely by the clinical team.

