## Supplementary material for "Placental LBX1 dysregulation drives ribosomal dysfunction and unfolded protein response in spontaneous preterm birth": LBX1_Supporting Information

^†^ Both authors contribute equally to this work

^‡^Study Cohort

Corresponding authors:

Tushar Kanti Maiti:

Pallavi Kshetrapal:

**Note:** The supporting information includes Supplementary File S1-S7 attached as excel and Supplementary File S8 attached as PDF format.

**Supporting Information – Table of Contents**

| **Index** | **Title** | **Page No.** |
| --- | --- | --- |
| **Supplementary Figure S1** | Flow-charts for clinical sample selection | **S4** |
| **Supplementary Figure S2** | Proteome data quality, sub-cellular localization and cell-type abundance in placenta samples | **S5-S6** |
| **Supplementary Figure S3** | Construction and evaluation of protein co-expression networks | **S7-S8** |
| **Supplementary Figure S4** | The quantitative prediction of hub-proteins and their expression trend across the period of gestation. | **S9-S10** |
| **Supplementary Figure S5** | The predictability and abundance trajectory of top performing proteins and data quality of MRM runs | **S10-S11** |
| **Supplementary Figure S6** | The LBX1 modulated proteome data quality assessment and its functional dynamics. | **S12-S13** |
| **Supplementary Figure S7** | Cluster 2 validation, ribosomal protein regulation, LBX1 IP data quality and true interacting protein enriched pathways | **S14-S15** |
| **Supplementary Figure S8** | The densitometric quantification of ribosomal homeostasis related targets | **S15-S16** |
| **Supplementary File S1** | Placental cell-type markers and their calculated abundance in clinical placental samples | **Attached Excel file** |
| **Supplementary File S2** | Differentially expressed proteins and pathways in LPTB vs TB and EPTB vs TB comparison | **Attached Excel file** |
| **Supplementary File S3** | The proteins clustered in blue and yellow modules and their GO enriched pathways | **Attached Excel file** |
| **Supplementary File S4** | Data-matrix and differential analysis of HR-MRM acquired maternal plasma quantitation of 13 hub proteins | **Attached Excel file** |
| **Supplementary File S5** | Differentially regulated proteins in OE vs COE and KD vs CKD comparison | **Attached Excel file** |
| **Supplementary File S6** | The clusters of differential proteins from OE vs COE and KD vs CKD comparisons and their cluster specific pathway over-representation using GO database. | **Attached Excel file** |
| **Supplementary File S7** | The SAINTexpress mediated probabilistic scoring of identified proteins and GO pathway enrichment of true interacting proteins. | **Attached Excel file** |
| **Supplementary File S8** | Uncropped original blots | **Attached PDF file** |

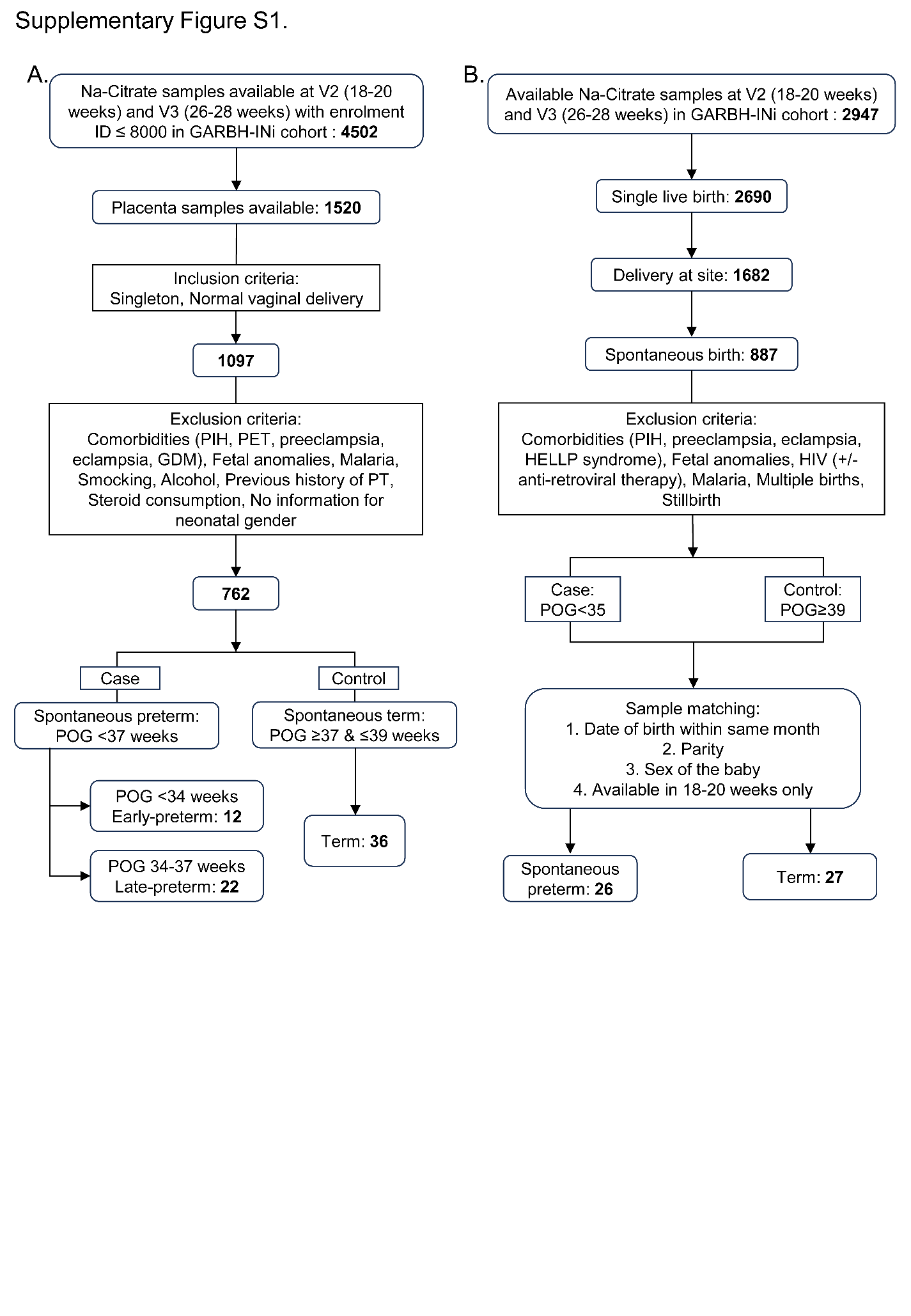

**Supplementary Figure S1:** Flow-charts for clinical sample selection **A)** A multi-stage cross-sectional study design is employed to select 70 participants following the flow-chart. **B)** The 18-20 weeks of maternal plasma samples (n=53) were selected according to the flow chart.

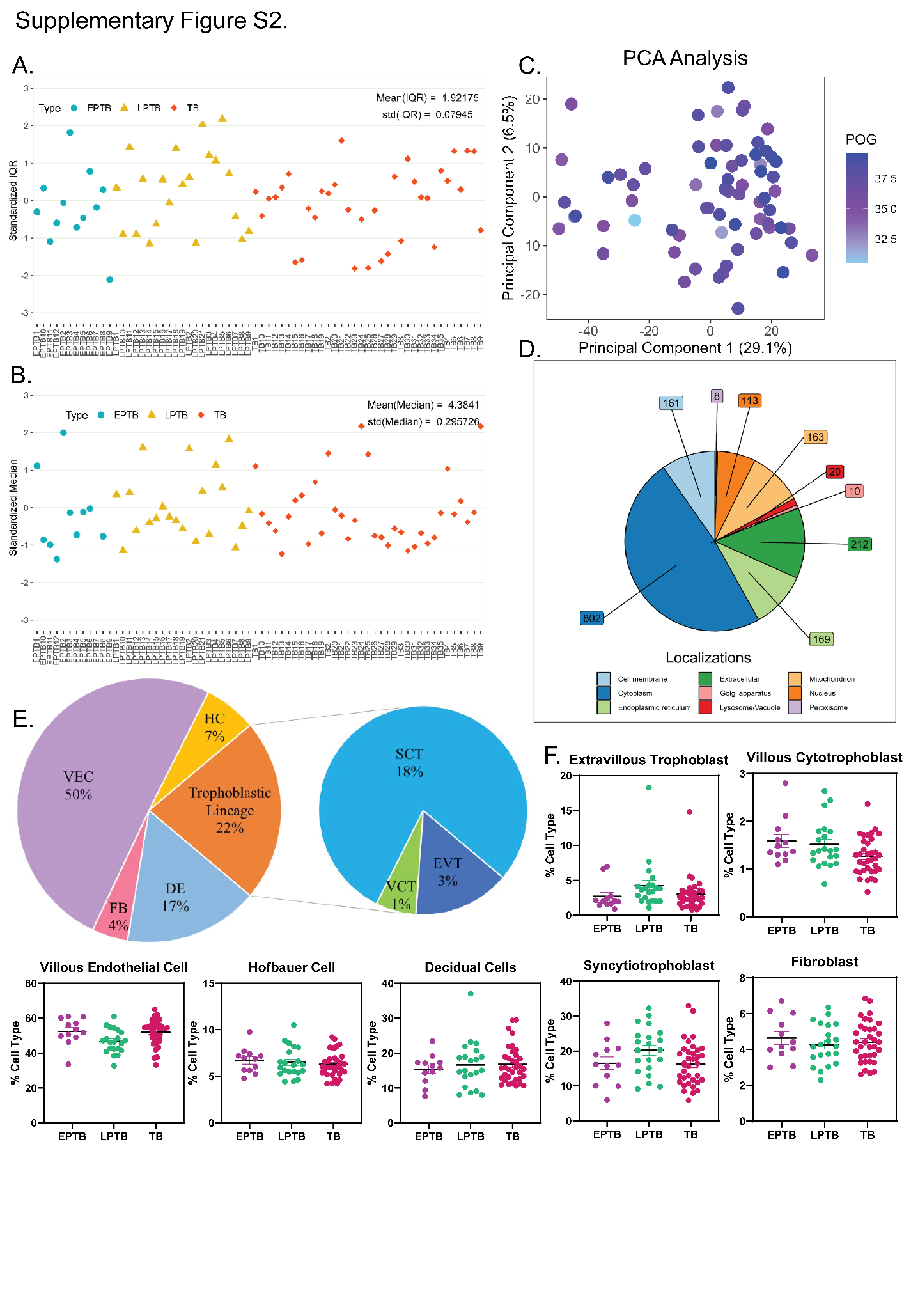

**Supplementary Figure S2:** Proteome data quality, sub-cellular localization and cell-type abundance in placenta samples. The plots show the spread of **A)** standardized IQR and **B)** standardized median across the 70 placental samples, with blue dots showing EPTB group, yellow triangle denoting LPTB group and orange kites as TB groups. **C)** The PCA clustering of placenta samples according to GA at delivery. PC1 and PC2 is plotted on x and y-axis respectively. **D)** The sub-cellular compartment allocation of placenta identified proteins are visualized with a pie chart. Each compartments are coloured differently and the compartment associated protein numbers are mentioned outside the circle. **E)** The pie-chart demonstrates the gross percentage distribution of different cell types from both trophoblastic and non-trophoblastic lineages. **F)** The percentage abundance of each cell types was plotted in a box-plot with the samples grouped according to the comparison condition.

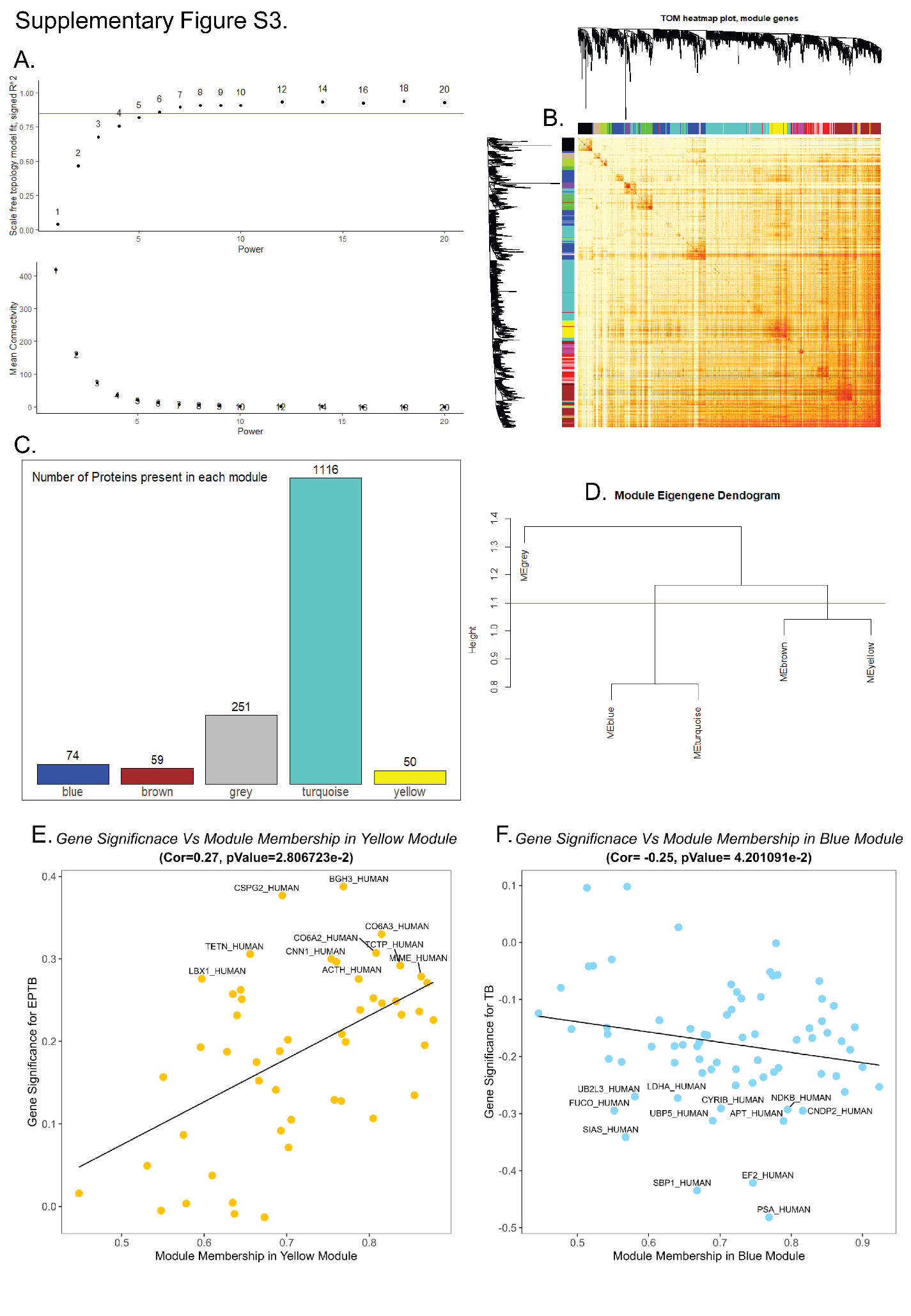

**Supplementary Figure S3:** Construction and evaluation of protein co-expression networks. **A)** Mean connectivity and scaled independence are calculated for increasing gradient of soft thresholds. **B)** The topological overlap matrix (TOM) derived degree of correlation between different unmerged coloured modules. The gradient of light to dark colours indicates the lower to higher correlation pattern. **C)** The proteins in each coloured modules were reported using bar-plot. **D)** The hierarchical clustering dendrogram between the coloured modules indicates how close the modules are to each other. The scatter-plot visualized with module membership on x-axis and gene significance on y-axis for **E)** yellow and **F)** blue module indicates its correlative trend and relevance of proteins in each cluster.

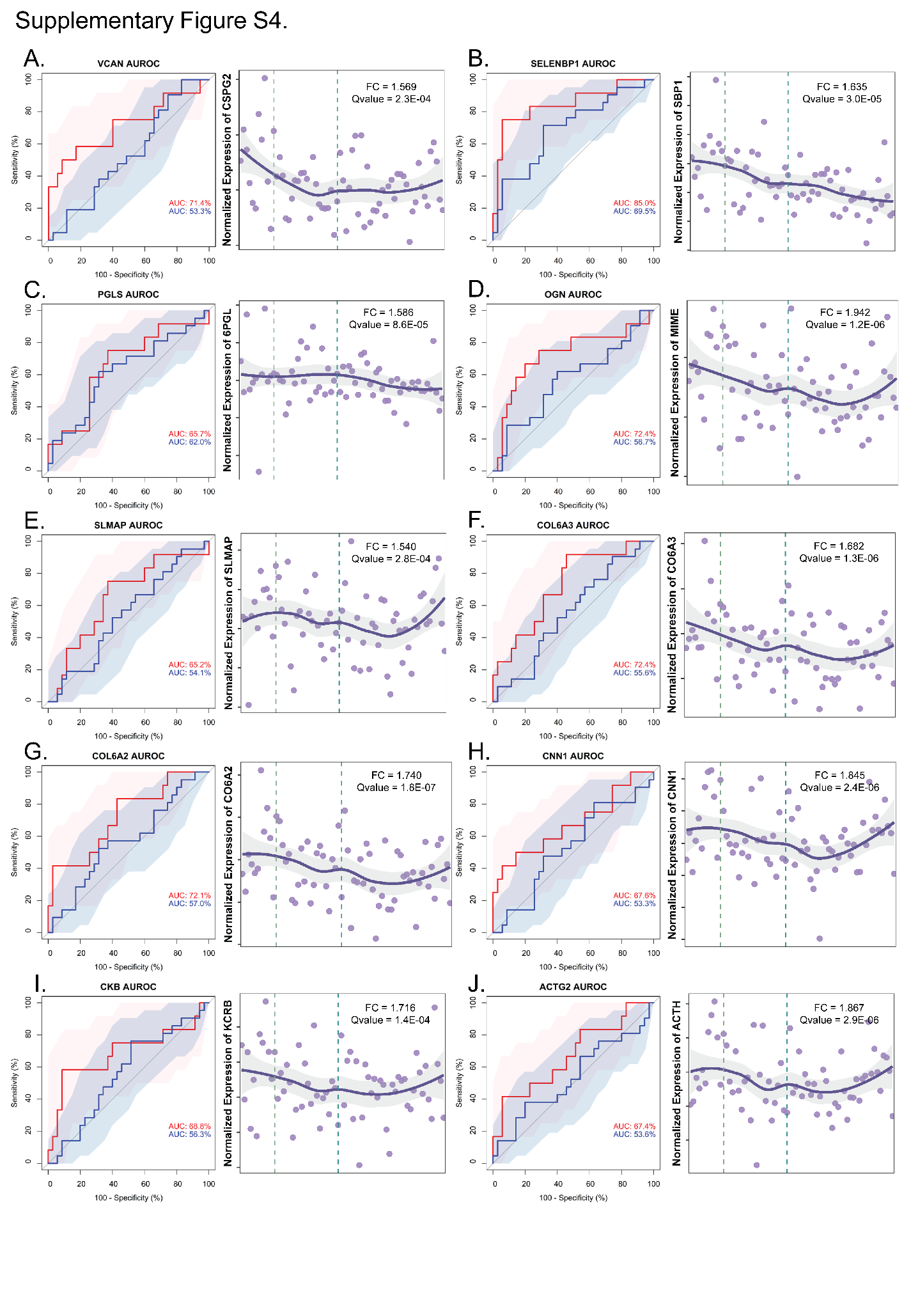

**Supplementary Figure S4:** The quantitative prediction of hub-proteins and their expression trend across the period of gestation. The AU-ROC is measured and the longitudinal expression trend is plotted for **A)** VCAN, **B)** SELENBP1, **C)** PGLS, **D)** OGN, **E)** SLMAP, **F)** COL6A3, **G)** COL6A2, **H)** CNN1, **I)** CKB and **J)** ACTG2.

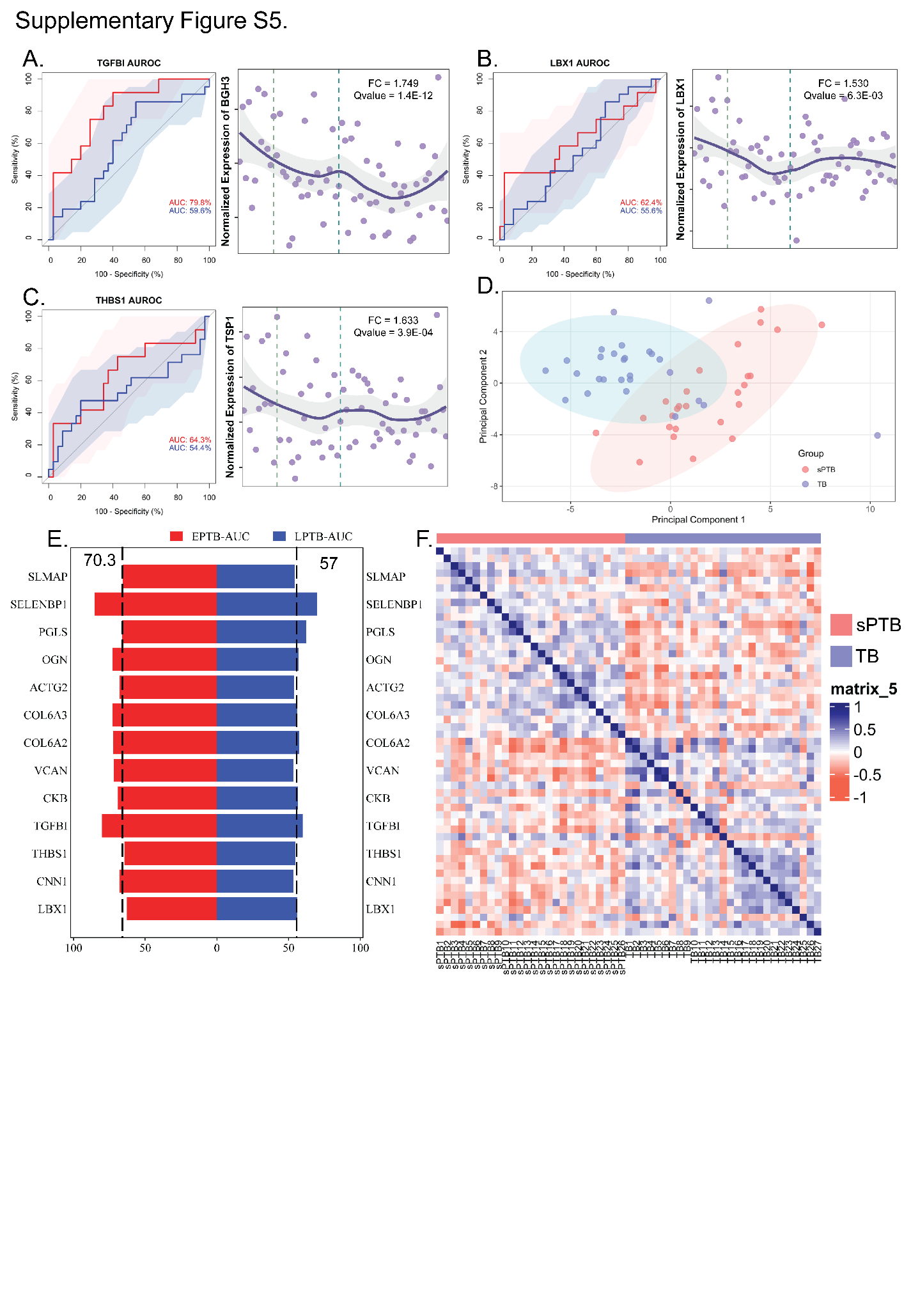

**Supplementary Figure S5:** The predictability and abundance trajectory of top performing proteins and data quality of MRM runs. The AU-ROC and profile trend across POG is plotted for **K)** TGFBI, **L)** LBX1 and **M)**THBS1. **N)** The calculated AU-ROC predicting EPTB and LPTB outcome for each hub-protein is tabulated in a bar diagram. The red indicates EPTB prediction and blue indicate LPTB prediction. The MRM acquired plasma quantified hub protein intensity matrix is able to group the sPTB samples from TB samples visualized using the **O)** PCA clustering and **P)** heatmap of Pearson’s correlation coefficient.

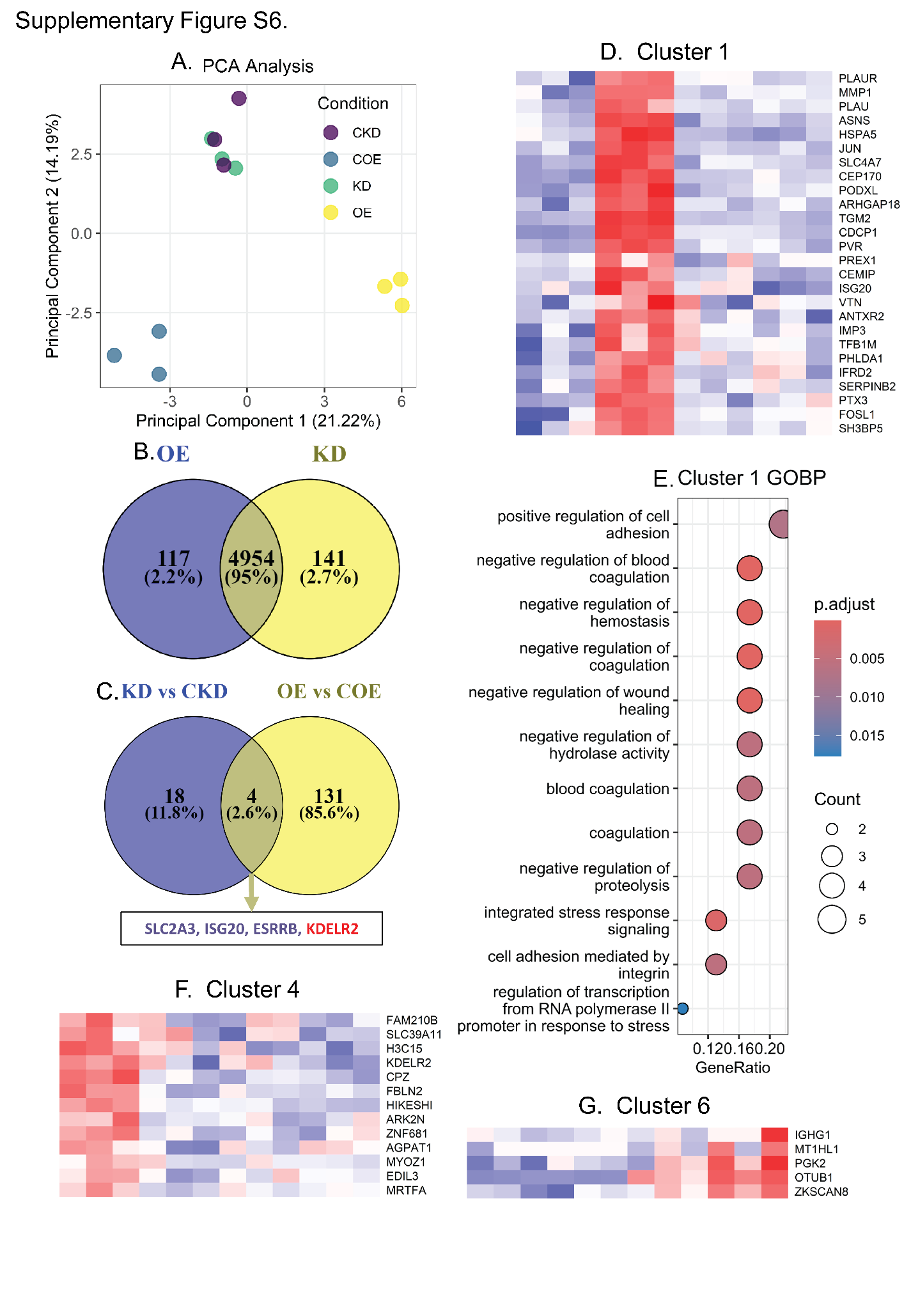

**Supplementary Figure S6:** The LBX1 modulated proteome data quality assessment and its functional dynamics. **A)** The PCA clustering of over-expression (OE, COE) and knock down (KD, CKD) related samples with PC1 plotted on x-axis and PC2 plotted on y-axis. The Venn diagram demonstrating the overlap between **B)** identified and **C)** differentially expressed proteins (DEPs). The common DEPs are mentioned in the square below. **D)** The cluster 1 grouped proteins and their expression dynamics across the samples. **E)** Cluster 1 proteins enriched GO pathways, where the circle size and colour indicate pathway associated proteins count and adjusted p-value respectively. The expression pattern of proteins clustered in **F)** cluster 4 and **G)** cluster 6.

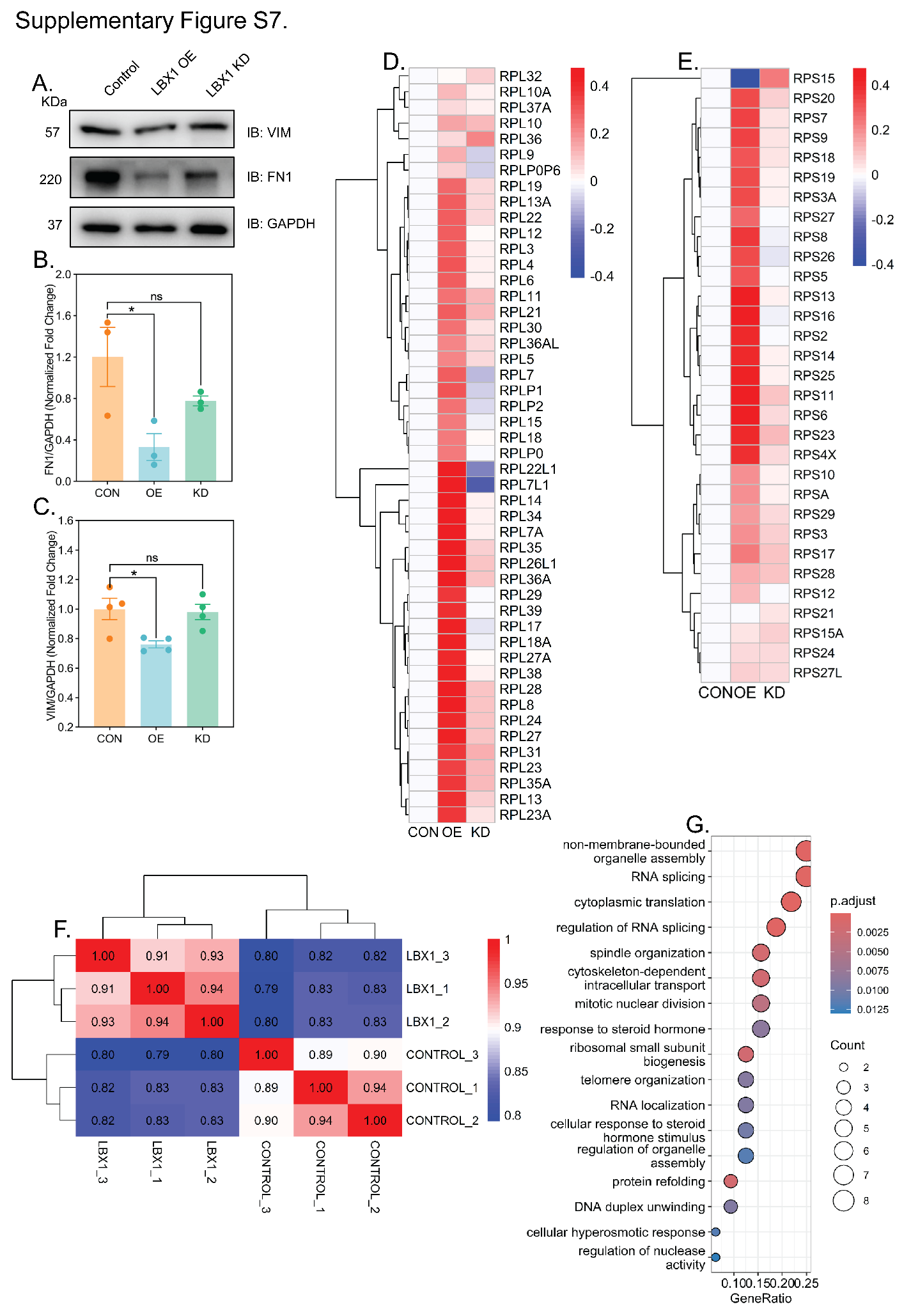

**Supplementary Figure S7:** Cluster 2 validation, ribosomal protein regulation, LBX1 IP data quality and true interacting protein enriched pathways. **A)** The immune-blot analysis of EMT markers (VIM and FN1) in LBX1 overexpression and knock down samples. GAPDH is used as loading control. The densitometric quantitative values of **B)** FN1 and **C)** VIM bands post GAPDH normalization is plotted as mean ± SEM (n=3). The student’s t-test measured p-value of <0.05 is considered as statistically significant. The heatmap of FC from OE vs COE and KD vs CKD comparison were visualized for ribosomal **D)** large subunit and **E)** small subunit proteins. Red colour indicates higher FC, while blue indicates lower FC. **F)** The calculated Pearson’s correlation coefficient between samples of LBX1 IP and control were plotted in a heatmap, where the red and blue denote higher and lower correlation values respectively. **G)** The over-representation of GO pathways using SAINT value filtered proteins shows the pathways in y-axis, with the circle size indicating the number of associated proteins and the circle colour denoting adjusted p-values for respective pathways.

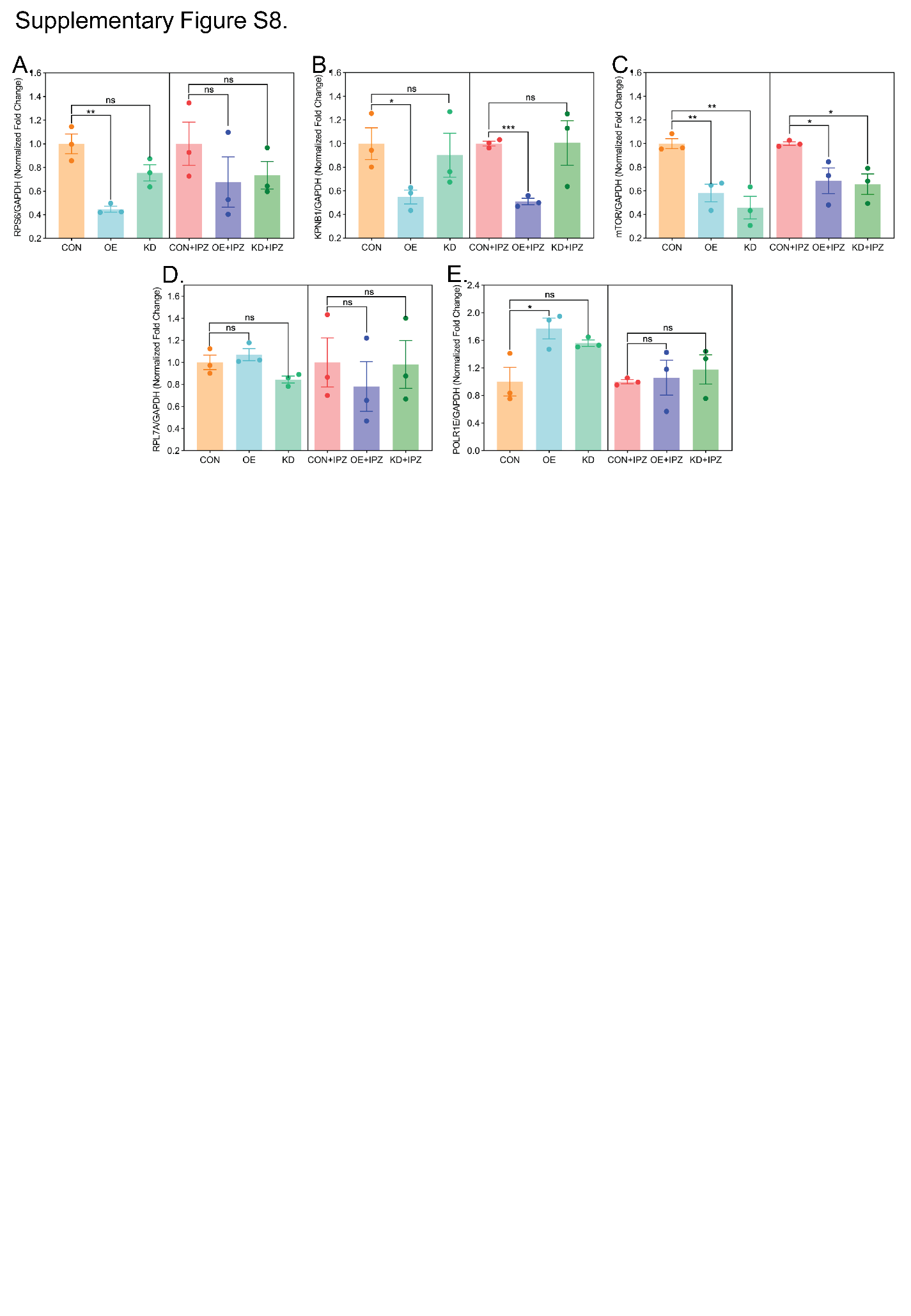

**Supplementary Figure S8:** The densitometric quantification of ribosomal homeostasis related targets. The protein band intensity of **A)** RPS6, **B)** KPNB1, **C)** mTOR, **D)** RPL7A and, **E)** POLR1E are quantified using ImageJ. The intensities from the three bio-replicates were further normalized using GAPDH intensity and then plotted as mean ± SEM. The significance was calculated using Student’s t-test where the p-value of <0.05 is considered statistically significant.
